# The relationship of maternal gestational mass spectrometry-derived metabolites with offspring congenital heart disease: results from multivariable and Mendelian randomization analyses

**DOI:** 10.1101/2022.02.04.22270425

**Authors:** Kurt Taylor, Nancy McBride, Jian Zhao, Sam Oddie, Rafaq Azad, John Wright, Ole A. Andreassen, Isobel D Stewart, Claudia Langenberg, Maria Magnus, Maria Carolina Borges, Massimo Caputo, Deborah A Lawlor

**Author notes:** Corresponding author – Oakfield House, MRC Integrative Epidemiology Unit Bristol, Population Health Science Institute, University of Bristol, BS8 2BN.

## Abstract

**Background:** It is plausible that maternal pregnancy metabolism influences risk of offspring congenital heart disease (CHD). We sought to explore this through a systematic approach using different methods and data.

**Methods:** We undertook multivariable logistic regression of the odds of CHD for 923 Mass Spectrometry (MS)-derived metabolites in a sub-sample of a UK birth cohort (Born in Bradford (BiB); N = 2,605, 46 CHD cases). We considered metabolites reaching a p-value threshold <0.05 to be suggestively associated with CHD. We sought validation of our findings, by repeating the multivariable regression analysis within the BiB cohort for any metabolite that was measured by nuclear magnetic resonance (NMR) or clinical chemistry (N = 7,296, 87 CHD cases), and by using genetic risk scores (GRS: weighted genetic risk scores of single nucleotide polymorphisms (SNPs) that were associated with each metabolite) in Mendelian randomization (MR) analyses. MR analyses were performed in BiB and two additional European birth cohorts (N = 38,662, 319 CHD cases).

**Results:** In the main multivariable analyses, we identified 44 metabolites suggestively associated with CHD, including those from the following super pathways: amino acids, lipids, co-factors and vitamins, xenobiotics, nucleotides, energy, and several unknown molecules. Of these 44, isoleucine and leucine were available in the larger BiB cohort (NMR), and for these the results were validated. MR analyses were possible for 27/44 metabolites and for 11 there was consistency with multivariable regression results.

**Conclusions:** In summary, we have used complimentary data sources and statistical techniques to construct layers of evidence. We found that amino acid metabolism during pregnancy, several lipids (more specifically androgenic steroids), and levels of succinylcarnitine could be important contributing factors for CHD.

## Introduction

Congenital heart diseases (CHDs) are the most common congenital anomaly affecting approximately 6-8 per 1000 live births and 10% of stillbirths. They are the leading cause of death from congenital anomalies ^1^. Approximately 20% of CHD cases can be attributed to known chromosomal anomalies, gene disorders or teratogens ^2^. The causes of the remaining cases are unknown. Identifying causes of CHDs is important for improving aetiological understanding and developing potential targets for intervention.

Metabolomics technologies have enabled the quantification of a large number of metabolites in a biological sample. Metabolites are small-molecule intermediates and products of metabolism. The metabolome, the complete set of metabolites in biological tissues/fluids, is influenced by both genotype and environment, and dynamically responds to environmental influences. Analyses of maternal metabolomic profiles could identify causal mechanisms leading to CHDs ^3^. Because the metabolome reflects interactions of genomic, environmental (e.g., air pollution), behavioural (e.g., smoking) and pathophysiological states (e.g., body composition), examining associations of it with CHDs could help elucidate modifiable upstream risk factors and/or potential molecular targets for intervention to prevent CHDs.

Studies have explored maternal molecular markers and found that offspring of women with a compromised vitamin D status (defined as 25-hydroxyvitamin D < 50 nmol/l in comparison to adequate defined as > 75 nmol/l) ^4^ and lipid profile ^5,6^ have an increased risk of CHDs. Other work has shown that poor glucose control and diabetes during pregnancy can increase CHD risk ^7–9^. However, these studies focus on single or few biomarkers. Exploring the wider metabolome could provide opportunities to improve our understanding of the molecular mechanisms that underpin CHDs ^3^. Previous work has explored metabolomics in maternal serum as a predictor of offspring CHDs and uncovered potentially relevant biological pathways ^10^. The study found more than 100 metabolites that differed between CHD cases and non-cases concluding that abnormal lipid metabolism was an important feature of CHD pregnancies. Other research has explored potential biomarkers of maternal urine metabolomics with offspring CHDs (N = 70 CHD cases and 70 controls) ^11^. Their results indicated that short chain fatty acids and aromatic amino acid metabolism may be relevant to CHDs. Replication of these results are warranted. A recent retrospective study in a Chinese population performed metabolomic analyses using maternal amniotic fluid and found that two metabolites (uric acid and proline) were elevated in CHD affected pregnancies ^12^. In summary, there have been studies uncovering potentially important biological pathways associated with offspring CHDs. However, pregnancy metabolomic studies are still relatively novel with scope for future research to provide new insights and seek replication of previous findings.

The aim of this study was to explore associations of the maternal metabolome quantified by an untargeted mass spectrometry (MS) platform and the odds of CHD in the offspring. To address this aim we searched for relevant studies within The LifeCycle Project-EU Child Cohort Network ^13^ to identify any study with detailed untargeted maternal gestational metabolomic data and offspring CHD information. We identified only one cohort with relevant data in a subgroup: the Born in Bradford (BiB) cohort (N = 2,605 participants; 46 CHD cases) ^14,15^. Recognising that these novel data were potentially underpowered, we sought internal validation of metabolites suggestively associated with CHD, by repeating the multivariable regression analysis within the BiB cohort for any metabolite that was measured by nuclear magnetic resonance (NMR) or clinical chemistry in larger numbers (N = 7,296, 87 CHD cases). We subsequently searched the MR-PREG consortia studies ^16,17^ for cohorts with maternal genome-wide data and offspring CHD information that could be used for Mendelian randomization (MR) analyses of associations of genetic instruments for maternal metabolites. We performed pooled MR analyses across three MR-PREG cohorts meeting our criteria (N = 38,663, 319 CHD cases) for any metabolites that were: (i) suggestively associated with CHD in BiB (P<0.05) and (ii) had summary data in the most recent metabolomic genome-wide association study (GWAS).

## Methods

### Study design and participants

A schematic overview of the study design is illustrated in **Figure 1**. We excluded children of multiple births because they differ from single births for congenital anomaly outcomes ^18,19^. For multivariable metabolomic analyses, we used data from the BiB cohort as this was the only cohort that had measures of a substantial number of metabolites reflecting a wide range of metabolic paths assessed during pregnancy and CHD outcomes ^15^. We also explored internal validation of any findings with a p-value < 0.05 within the BiB study where equivalent (or near equivalent) measures to any on the MS platform markers are available from other sources. BiB is a population-based prospective birth cohort, including 12,453 women across 13,776 pregnancies who were recruited at their oral glucose tolerance test (OGTT) at approximately 26–28 weeks’ gestation ^14^. Eligible women had an expected delivery between March 2007 and December 2010. The use of a multivariable p-value threshold of <0.05 to take associations forward into further validation analyses is appropriate as an initial screen, for a relatively rare outcome, to avoid missing potential causal effects.

**Figure 1.**
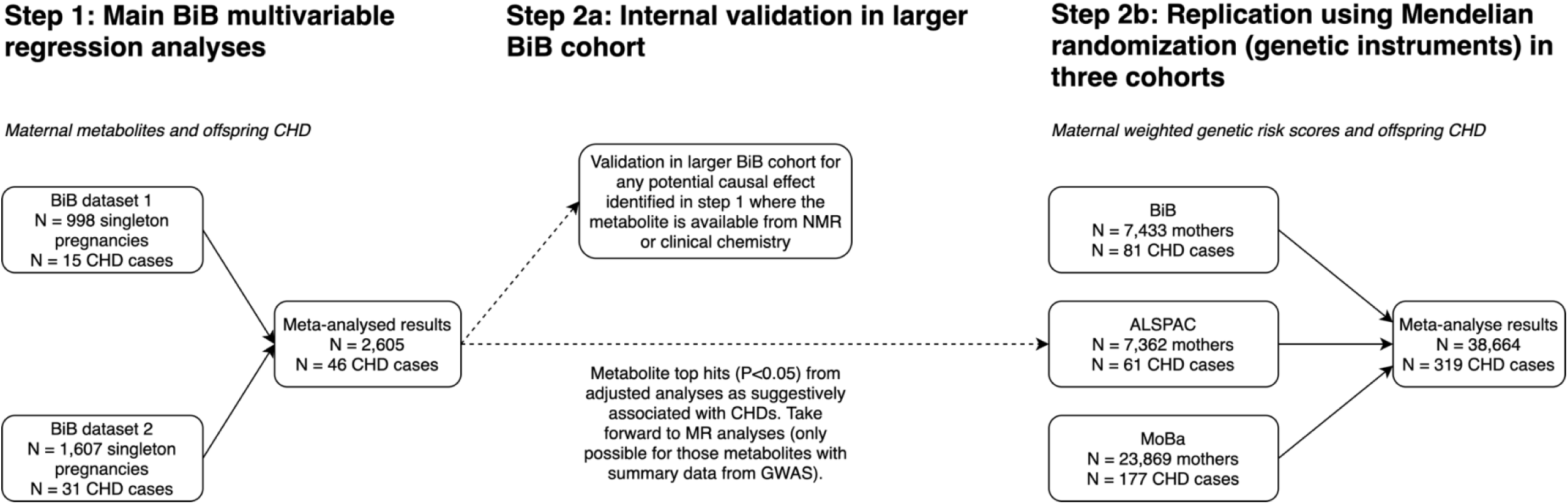
An overview of the study design. BiB has pregnancy mass spectrometry derived metabolomics in two separate datasets. Dataset 1 was completed in December 2017 and included 1,000 maternal pregnancy samples. Dataset 2 was completed in December 2018 and consisted of 2,000 maternal pregnancy samples within a case cohort design. The selection of participants into the two MS metabolomic datasets are shown in flowcharts in Figure S1. Abbreviations: CHD, congenital heart disease; BiB, Born in Bradford; NMR, Nuclear Magnetic Resonance; MR, Mendelian Randomization; GWAS, genome-wise association study; ALSPAC, Avon Longitudinal Study of Parents and Children; MoBa, Norwegian Mother, Father and Child Cohort.

To be included in MR analyses, studies and participants had to have genome-wide data in mothers and CHD data in the offspring. Three cohorts contributed to MR analyses: BiB, the Avon Longitudinal Study of Parents and Children (ALSPAC) and the Norwegian Mother, Father and Child Cohort Study (MoBa). ALSPAC is a UK prospective birth cohort study which was devised to investigate the environmental and genetic factors of health and development ^20–22^. Pregnant women resident in Avon, UK with expected dates of delivery 1st April 1991 to 31st December 1992 were invited to take part in the study. The initial number of pregnancies enrolled is 14,541 (for these at least one questionnaire has been returned or a “Children in Focus” clinic had been attended by 19/07/99). Of these initial pregnancies, there was a total of 14,676 fetuses, resulting in 14,062 live births and 13,988 children who were alive at 1 year of age. MoBa is a population-based pregnancy cohort study conducted by the Norwegian Institute of Public Health ^23,24^. Participants were recruited from all over Norway from 1999-2008. The women consented to participation in 41% of the pregnancies. The cohort includes approximately 114,500 children, 95,200 mothers and 75,200 fathers. The current study is based on 12 of the quality-assured data files released for research in 2019. The establishment of MoBa and initial data collection was based on a license from the Norwegian Data Protection Agency and approval from The Regional Committees for Medical and Health Research Ethics. The MoBa cohort is currently regulated by the Norwegian Health Registry Act. The current study was approved by The Regional Committees for Medical and Health Research Ethics (2018/1256).

### Sample collection and metabolomic profiling in BiB

Of the 13,776 pregnancies in the BiB cohort, 11,480 had a fasting blood sample taken during the OGTT (n = 10,574 [92%] between 26–28 weeks’ gestation, with the remaining women being within 11–39 weeks’ gestation). Samples were taken by trained phlebotomists working in the antenatal clinic of the Bradford Royal Infirmary and sent immediately to the hospital laboratory. The metabolomics data in the BiB cohort has previously been described in detail ^15^. In brief, metabolomics analysis was performed on ethylenediamine tetraacetic acid (EDTA) plasma samples around 26-28 weeks’ gestation. The untargeted MS metabolomics analysis of over 1,000 metabolites was performed at Metabolon, Inc. (Durham, North Carolina, USA). Quality control of the metabolite data was conducted by Metabolon. The classes of metabolites include amino acids, carbohydrates, cofactors and vitamins, energy, lipids, nucleotides, partially characterised molecules, peptides, and xenobiotics. These super-pathways, as defined by Metabolon, are also further subdivided into ∼80 sub-pathways. Metabolite concentrations were quantified using area under the curve of primary MS ions and were expressed as the multiple of the median (MoM) value for all batches processed on the given day. The MoM more closely reflects the biological variation rather than technical variation between samples or analysis platform ^25^. Due to the timing of funding acquisition, samples were sent to Metabolon in two separate batches. Dataset 1 was completed in December 2017 and included 1,000 maternal pregnancy samples. Dataset 2 was completed in December 2018 and consisted of 2,000 maternal pregnancy samples within a case cohort design. Over-sampled cases were removed to obtain a representative sample. The selection of participants into the two MS metabolomic datasets are shown in flowcharts in Figure S1 and have been described in detail previously ^15^.

### Confounders

In multivariable regression analyses in BiB, we adjusted for the following maternal characteristics based on their known or plausible influence on maternal metabolites and on CHD: age, ethnicity, parity, residential neighbourhood Index of Multiple Deprivation (IMD), body mass index (BMI), smoking, and alcohol consumption. Details of the methods for how confounders were assessed are provided in the Supplementary File 1 (Text S1).

### Congenital heart disease outcomes

In BiB, cases were identified from either the Yorkshire and Humber congenital anomaly register database, which will tend to pick up most cases that were diagnosed antenatally and in the early postnatal period of life, or through linkage to primary care (up until aged 5), which will have picked up any additional cases, in particular those that might have been less severe and not identified antenatally/in early life ^26^. All BiB cases were confirmed postnatally and were assigned ICD-10 codes. We used ICD-10 codes to assign CHD cases according to the European surveillance of congenital anomalies (EUROCAT) guidelines. In the ALSPAC cohort, cases were obtained from a range of data sources, including health record linkage and questionnaire data up until age 25 following European EUROCAT guidelines ^27^. In MoBa, information on whether a child had a CHD or not (yes/no) was obtained through linkage to the Medical Birth Registry of Norway (MBRN). All maternity units in Norway must notify births to the MBRN, and information on malformations are reported to the registry up to 12 months postpartum ^28^. Further details on defining CHDs including ICD codes are shown in Text S2 and Table S1 (Supplementary File 1).

### Genetic data

The rationale for performing MR analyses was to explore replication using a different method with two additional independent studies and to explore causation. Metabolites are affected by multiple disease processes as well as numerous environmental exposures; therefore, understanding the metabolic pathways implicated in CHD is nontrivial. MR can help discriminate causal from non-causal metabolites because genetic variants are less likely to be confounded by the socioeconomic and environmental factors that might bias causal estimates in conventional multivariable regression ^29^, but may be biased by a path from the metabolomic genetic score to CHD, for example via horizontal pleiotropy or fetal genotype ^30^. Consistent results from both increase confidence that the result is causal.

#### Genotyping in each cohort

ALSPAC mothers were genotyped using Illumina human660K quad single nucleotide polymorphism (SNP) chip, and ALSPAC children were genotyped using Illumina HumanHap550 quad genome-wide SNP genotyping platform. Genotype data for both ALSPAC mothers and children were imputed against the Haplotype Reference Consortium v1.1 reference panel, after performing the QC procedure (minor allele frequency (MAF) ≥ 1%, a call rate ≥ 95%, in Hardy-Weinberg equilibrium (HWE), correct sex assignment, no evidence of cryptic relatedness, and of European descent). The samples of the BiB cohort (mothers and offspring) were processed on three different type of Illumina chips: HumanCoreExome12v1.0, HumanCoreExome12v1.1 and HumanCoreExome24v1.0. Genotype data were imputed against UK10K + 1000 Genomes reference panel, after a similar QC procedure (a call rate ≥ 99.5%, correct sex assignment, no evidence of cryptic relatedness, correct ethnicity assignment). In MoBa, blood samples were obtained from both parents during pregnancy and from mothers and children (umbilical cord) at birth ^31^. Genotyping has had to rely on several projects - each contributing with resources to genotype subsets of MoBa over the last decade. The data used in the present study was derived from a cohort of genotypes samples from four MoBa batches. The MoBa genetics QC procedure involved MAF ≥ 1%, a call rate ≥ 95%, in HWE, correct sex assignment, and no evidence of cryptic relatedness. Further details of the genotyping methods for each cohort are provided in the Supplementary File 1 (Text S3) including flow charts showing selection of participants (Figure S2).

#### GWAS data and SNP selection

We aimed to construct weighted GRSs for metabolites that had a p-value <0.05 (referred to throughout as “suggestively associated” with CHDs) in the multivariable regression analyses using BiB data. To do this, we cross-referenced our suggestive associations with large relevant GWAS. We used summary data from two GWAS. In the first, the authors explored the genetic effects of 174 metabolites (compared with the 923 included in our study) ^32^. To ensure independent associations, SNPs used from the first GWAS were selected at p < 5 × 10^−8^ and were clumped to ensure independence at linkage disequilibrium (LD) r^2^ = 0.001 and a distance of 10,000 kb using the TwoSampleMR package ^33^. In the second (unpublished), the authors performed a GWAS of metabolon metabolite levels using samples from the EPIC-Norfolk ^34^ and INTERVAL studies ^35^. 14,296 participants were included in a discovery set (5,841 from EPIC-Norfolk; 8,455 from INTERVAL) and 5,698 from EPIC-Norfolk in a validation set. The authors performed exact conditional analyses to identify independent associations. A total of 913 metabolites were taken forward for their GWAS analysis.

#### Genetic risk score generation

GRSs were calculated using SNPs previously associated in largescale GWAS with metabolites (described above) by adding up the number of metabolite increasing alleles among the selected SNPs after weighting each SNP by its effect on the corresponding metabolite:

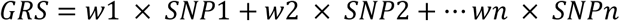

where w is the weight (i.e., the beta-coefficient of association of the SNP with the exposure from the published GWAS) and SNP is the genotype dosage of exposure-increasing alleles at that locus (i.e., 0, 1, or 2 exposure-raising alleles). After matching metabolites suggestively associated with CHDs at P<0.05 from multivable regression analyses and removing indels, selected SNPs were extracted from the imputed genotype data in dosage format using QCTOOL (v2.0) and VCF tools (v 0.1.12b) in ALSPAC and BiB, respectively. PLINK (v1.9) was then used to construct the GRS for each exposure coded so that an increased score associated with increased levels of metabolite. In MoBa, we constructed the GRSs from the QC’d data in PLINK format. If a SNP was missing, a proxy SNP was used where available based on r^2^ > 0.8 using the European reference panel in the LDLink R package ^36^.

### Statistical analysis

Analyses were performed in R version 4.0.2 (R Foundation for Statistical Computing, Vienna, Austria). An analysis plan was written and uploaded to the Open Science Framework before analyses commenced, where any subsequent changes to analyses were documented along with the rationale ^37^. We used scaled imputed data (in which missing data have been imputed and the multiple of median values transformed to standard deviation (SD)-scores) which was log transformed. Any metabolite (in either dataset) where there was too little variation for meaningful analyses (defined as < 440 unique values) was excluded ^38^. Transformed metabolite values were converted to standard deviation SD units. There were 1,100 and 1,150 quantified metabolites included in dataset 1 and 2, respectively, with 923 of these present in both datasets.

#### Multivariable regression (metabolomic) analyses

We used logistic regression to estimate odds ratios (ORs) and 95% confidence intervals (CIs) of any CHD per SD higher metabolite, with and without adjustment for confounders. As we are interested in potential causal effects, we present confounder adjusted results throughout. Analyses were done separately in the two BiB datasets and results pooled using fixed-effects meta-analyses. Given that CHD is rare and binary, we accepted an uncorrected P<0.05 (from meta-analyses) for the metabolite being suggestively associated with CHD in the offspring (but requiring further validation). We took these metabolites forward to MR analyses.

The MS-platform used in BiB includes measures of xenobiotics which are synthetic chemicals that are not synthesised by humans. Their presence in the circulation usually reflects endogenous exposures, such as medications and supplements. Given that these metabolites would not be present in all participants (and therefore have high missingness), many were removed (86/154 (56%)) from the dataset given the metabolite inclusion criteria of 440 unique observations mentioned above. Therefore, we performed an exploratory additional analysis using xenobiotics (N = 154) as binary variables (1 = yes; metabolite is detected in the sample, 0 = no; metabolite is not detected in the sample). We present adjusted ORs of these binary variables (any presence vs none) with CHDs.

We sought to internally validate any of the metabolites suggestively associated with CHDs that were also measured in BiB in larger numbers using different methods. After matching suggestive associations, we used data from the NMR platform (N = 2 metabolites) and did not use any data from the clinical chemistry measurements. More information on the BiB NMR data including methods, QC and participant information has been described in detail previously ^15^.

#### Mendelian randomization analyses

We undertook MR in each of the 3 cohorts, including all BiB, ALSPAC, and MoBa participants with maternal genetic data and offspring CHD data. Logistic regression was used to estimate the OR of CHD per SD change in GRS, with adjustment for the first 10 genetic principal components (PCs) with additional adjustment for genetic chip, genetic batch, and imputation batch in MoBa.

The key assumptions for MR are: (i) relevance assumption - the genetic instruments are robustly associated with the exposure and relevant to the population being studied (i.e. here pregnant women). We tested the association of the GRS of each metabolite with metabolite levels during pregnancy in BiB dataset 2. (ii) Independence assumption - The IV outcome association is not confounded. Such confounding could occur as a result of population stratification. To minimise this, we adjusted GRS-CHDs associations for the first 10 genetic PCs. We also repeated the MR analyses without the inclusion of BiB, given that BiB has a unique ethnic structure of South Asians and White Europeans. (iii) Exclusion restriction criteria - The genetic variant is not related to the outcome other than via its association with the exposure. We assessed pleiotropy by estimating the variance explained in all metabolites by each of the GRSs by undertaking the linear regression of every metabolite measured in BiB on each GRS. If the variance explained in other metabolites was similar or greater than to that explained in the candidate risk metabolite, this would suggest that there is low metabolite-specificity for the GRS and potential horizontal pleiotropic bias via the other metabolite(s). Importantly, however, this approach of testing GRS specificity does not distinguish between vertical pleiotropy (e.g. the GRS influences the candidate metabolite which is the precursor of another metabolite that affects CHD) and horizontal pleiotropy (e.g. the GRS influences two metabolites that affect CHD independently). We also check consistency of MR results when additionally adjusting for fetal genotype ^30^. We performed MR analyses separately in BiB, ALSPAC and MoBa and report pooled results from random-effect meta-analyses for all three cohorts and fixed-effect meta-analyses for MR analyses excluding BiB (i.e., ALSPAC and MoBa).

## Results

### Main BiB multivariable regression analyses

**Table 1** shows the distributions of characteristics for the women in both BiB datasets. In total, there were 2,605 mother-offspring pairs with 46 CHD cases included in the BiB multivariable regression metabolomic analyses. N.B. for consistency and clarity, we refer to metabolites here by their super-pathways (as defined by Metabolon). A metabolite might have a different super-pathway and chemical group. For example, N-Acetylcarnosine is a metabolite that is part of the amino acid super-pathway, but it is not an amino acid itself. Where available, we include Human Metabolome Database (HMDB) IDs with all numerical results which can assist the reader in finding further information on the structure and function of a metabolite. The super-pathways that included the largest proportions of the 923 metabolites were lipids (38%), unknown (22%), amino acids (18%) and Xenobiotics (8%), with other super-paths having ≤ 3% of the total (**Table 2)**.

**Table 1.** Participant characteristics for the Born in Bradford metabolomic analyses.

| Characteristic | Category | BiB dataset 1 (N = 998) | BiB dataset 2 (N = 1,607) |
| --- | --- | --- | --- |
| ***Offspring*** |  |  |  |
| CHD | Yes | 15 (1.6) | 31 (1.9) |
| Sex | Male | 510 (51.1) | 844 (52.5) |
|  | Female | 488 (48.9) | 763 (47.5) |
| ***Maternal*** |  |  |  |
| Age, years |  | 27.5 (5.7) | 27.3 (5.6) |
| Parity | Nulliparous | 358 (37.0) | 616 (36.8) |
|  | Multiparous | 610 (63.0) | 991 (63.2) |
| BMI, kg/m<sup>2</sup> |  | 26.7 (6.0) | 26.5 (5.8) |
| Ethnicity | White British | 500 (50.0) | 733 (45.6) |
|  | Pakistani | 498 (50.0) | 874 (54.4) |
| Neighbourhood deprivation (IMD) | Quintile 1 (most deprived) | 654 (65.5) | 1084 (67.5) |
|  | Quintile 2 | 175 (17.5) | 281 (17.5) |
|  | Quintile 3 | 112 (11.2) | 175 (10.9) |
|  | Quintile 4 | 38 (3.8) | 40 (2.5) |
|  | Quintile 5 (least deprived) | 19 (1.9) | 27 (2.7) |
| Smoking | Yes | 176 (17.7) | 311 (19.2) |
| Alcohol | Yes | 338 (33.9) | 496 (30.8) |
| Gest age at blood sampling, weeks |  | 26.2 (2.0) | 26.2 (2.0) |
| Data are means ± SD or n (%) unless stated. Abbreviations: BiB, Born in Bradford; CHD, congenital heart disease; BMI, body mass index; kg, kilogram; m, meter; HDP, hypertensive disorders of pregnancy; GHT, gestational hypertension; PE, pre-eclampsia; IMD, Index of Multiple Deprivation (taken from 2010 national quintiles); gest, gestational. |  |  |  |

**Table 2.** Showing the breakdown of metabolites in our dataset (N = 923) into the 10 super-pathways as defined by Metabolon.

| **Super pathway** | **N (%) for all metabolites (N = 923)** | **N (%) for metabolites suggestively associated with CHDs (N = 44)** |
| --- | --- | --- |
| Amino Acid | 170 (18.4%) | 10 (22.7%) |
| Lipid | 354 (38.4%) | 18 (40.9%) |
| Cofactors and Vitamins | 27 (2.9%) | 4 (9.0%) |
| Partially Characterized Molecules | 3 (0.3%) | 1 (2.3%) |
| Unknown | 201 (21.8%) | 7 (15.9%) |
| Xenobiotics | 86 (9.3%) | 2 (4.5%) |
| Nucleotide | 33 (3.6%) | 1 (2.3%) |
| Energy | 8 (0.9%) | 1 (2.3%) |
| Carbohydrate | 19 (2.1%) | 0 |
| Peptide | 22 (2.4%) | 0 |
| Abbreviations: CHD, congenital heart disease. |  |  |

Of the 923 metabolites quantified in both BiB datasets, 44 (4.8%) were associated with any CHD, at P < 0.05, in confounder adjusted pooled analyses (**Figure 2**). We observed suggestive effects (i.e., confounder adjusted associations reaching the p-value threshold <0.05) with several amino acids, lipids and co-factors and vitamins. There were also suggestive effects for two xenobiotics, one nucleotide, one energy metabolite and some partially characterised and unknown metabolites (**Figure 2**). None of the 22 peptide or 19 carbohydrate-related metabolites associated with CHD at this p-value threshold. Of the 18 lipid-related metabolites associated with CHD, 13 were positively associated (i.e., increased odds) (e.g., Glycolithocholate Sulfate: adjusted odds ratio (aOR) per SD increase in metabolite: 1.73 95% CI (1.21, 2.48)) and 5 were negatively associated (decreased odds) (e.g. Phosphocholine: aOR 0.65 (0.47, 0.90)). All but one (N−Acetylcarnosine) of the 10 amino acid-related metabolites were negatively associated with CHDs (e.g. isoleucine: aOR: 0.67 (0.49, 0.92)). 3 of the 4 co-factors and vitamins were negatively associated, whereas 1 (biliverdin) was positively associated (aOR 1.41 (1.07, 1.86)). The one nucleotide was negatively associated (inosine 5’−Monophosphate (Imp): aOR 0.59 (0.36, 0.99)) and the one energy related metabolite positively associated (succinylcarnitine (C4): aOR 1.42 (1.02, 1.97)). Benzoate and Saccharin were the two xenobiotics associated with CHDs in main analyses both showing positive associations. Results for associations of all metabolites (irrespective of p-value) in unadjusted and confounder adjusted analyses from the pooled datasets, and each dataset separately are provided in Supplementary File 2 (Tables S5-S7), including HMDB IDs where applicable.

**Figure 2.**
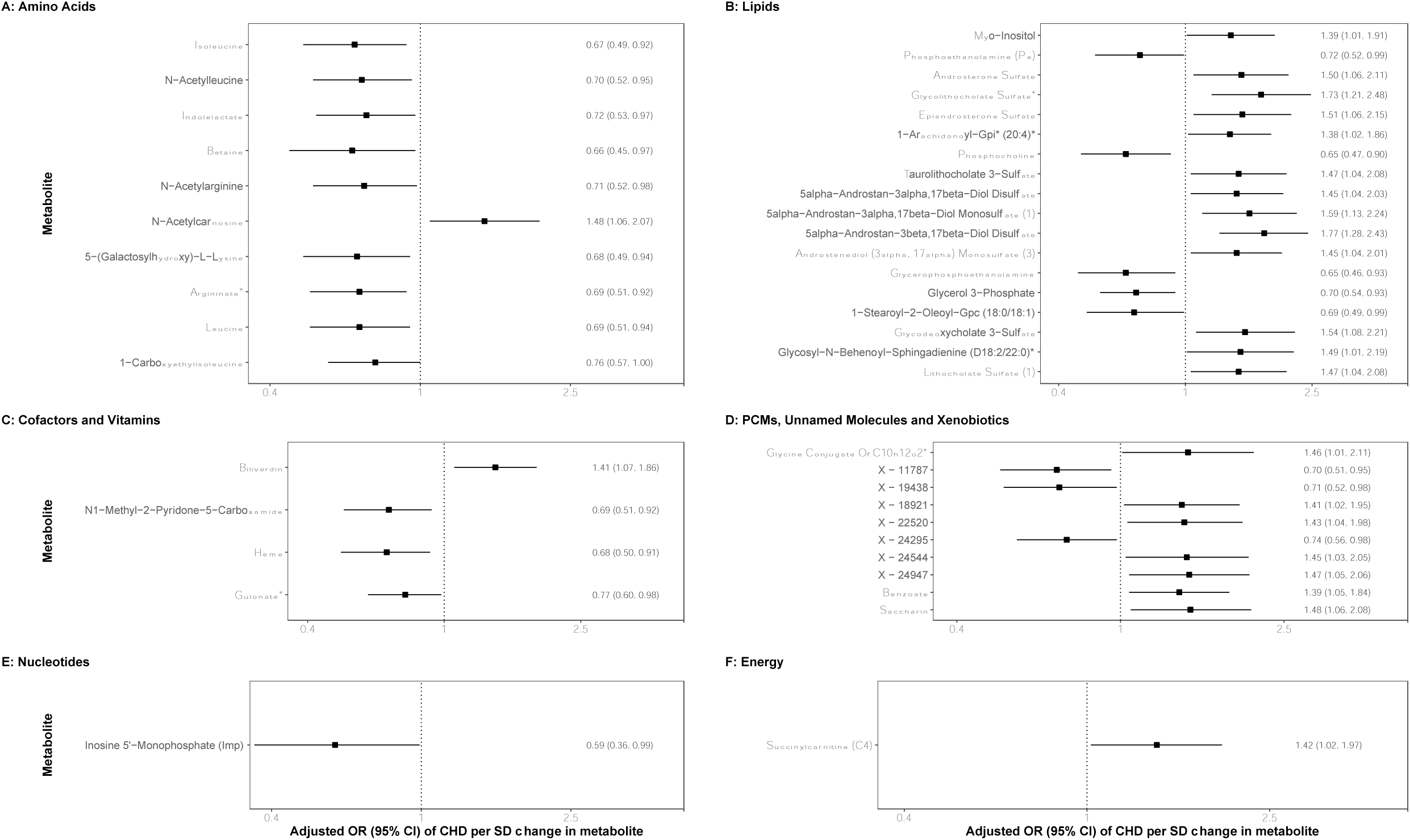
Pooled confounder adjusted associations of maternal pregnancy metabolites with offspring congenital heart disease in the Born in Bradford cohort (N = 2,391 & N CHD cases = 42) The associations show confounder adjusted odds ratios of CHD per standard deviation change in log-transformed metabolite levels for the 44 (out of 923) metabolites that associated with CHD at p-value <0.05 separated by super pathways as defined by Metabolon. Metabolites were measured at ∼26-28 weeks’ gestation. Heterogeneity statistics and separate associations for datasets 1 and 2 are reported in Supplementary Tables S5-S7. Associations were adjusted for maternal age, ethnicity, parity, Index of Multiple Deprivation, body mass index, smoking and alcohol intake. Abbreviations: PCMs, partially characterised molecules; OR, odds ratio; CHD, congenital heart disease; SD, standard deviation.

In the analysis treating xenobiotics as binary variables, after removal of metabolites with no exposed cases, there were 6 xenobiotic metabolites suggestively associated with offspring CHDs (Supplementary File 1: Table S2). 2 out of the 6 showed positive associations: saccharin, which was also associated in main analyses (adjusted odds ratio (aOR) for the presence of metabolite vs not: 2.16 95% CI (1.02, 5.13)) – an artificial sweetener) and salicyluric glucuronide (aOR: 2.27 (1.16, 4.29)) – a metabolite involved in aspirin metabolism). The remaining 4 showing negative associations are all part of the food component/plant metabolite sub pathway (Table S2).

### Internal validation using NMR or clinical chemistry measures of suggestive associations from main multivariable regression analyses

It was possible to explore 2 of the 44 metabolites suggestively associated with CHDs in the larger BiB sample. In comparable confounder adjusted analyses, NMR measured amino acids isoleucine and leucine were available on 7,296 mothers, with 87 having an offspring with CHD. Results for these two amino acids were highly consistent between the two samples/assay methods (aOR per SD increase in MS isoleucine 0.67 (0.49, 0.92) vs 0.65 (0.50, 0.84) for NMR isoleucine and aOR per SD increase in MS leucine 0.69 (0.51, 0.94) vs 0.67 (0.53, 0.85) for NMR leucine).

### Validating findings with Mendelian randomization

The distributions of offspring and maternal characteristics for MR analyses in BiB, ALSPAC and MoBa are displayed in Table S3 (Supplementary File 1). It was possible to explore MR replication for 27 of the 44 metabolites that associated with CHD in multivariable analyses (the other 17 were either not available in the GWAS or had genetic variants not available in the cohorts, Figure S3). All but 3 of the GRSs (24/27 (89%)) were associated with the corresponding metabolite during pregnancy in BiB (with R^2^ values ranging from 0.3% to 34%, for the remaining 3 the associations were wide with confidence intervals that included the null (Table S4; Supplementary File 1). Of the 27 GRSs, 3 were specific for the metabolite they were instrumenting (i.e. had the strongest association with it and little evidence of associations with other metabolites; N-acetylcarnosine, phosphocholine and succinylcarnitine). 18 GRSs were associated with the metabolite they were instrumenting and several others that were correlated with that metabolite (e.g. the biliverdin GRS was associated with it and also similarly with other hepatic-related metabolites). 6 GRSs were more strongly associated with other (uncorrelated) metabolites than the one they were instrumenting (scatter plots for all 27 GRSs are shown in Figure S4; Supplementary File 1). The 6 non-specific GRS were for indolelactate, glycolithocholate sulfate, isoleucine, leucine, myo-inositol and taurolithocholate 3-sulfate (MR results for these should be treated with caution and are denoted in **Figure 3B** by white-filled points).

**Figure 3.**
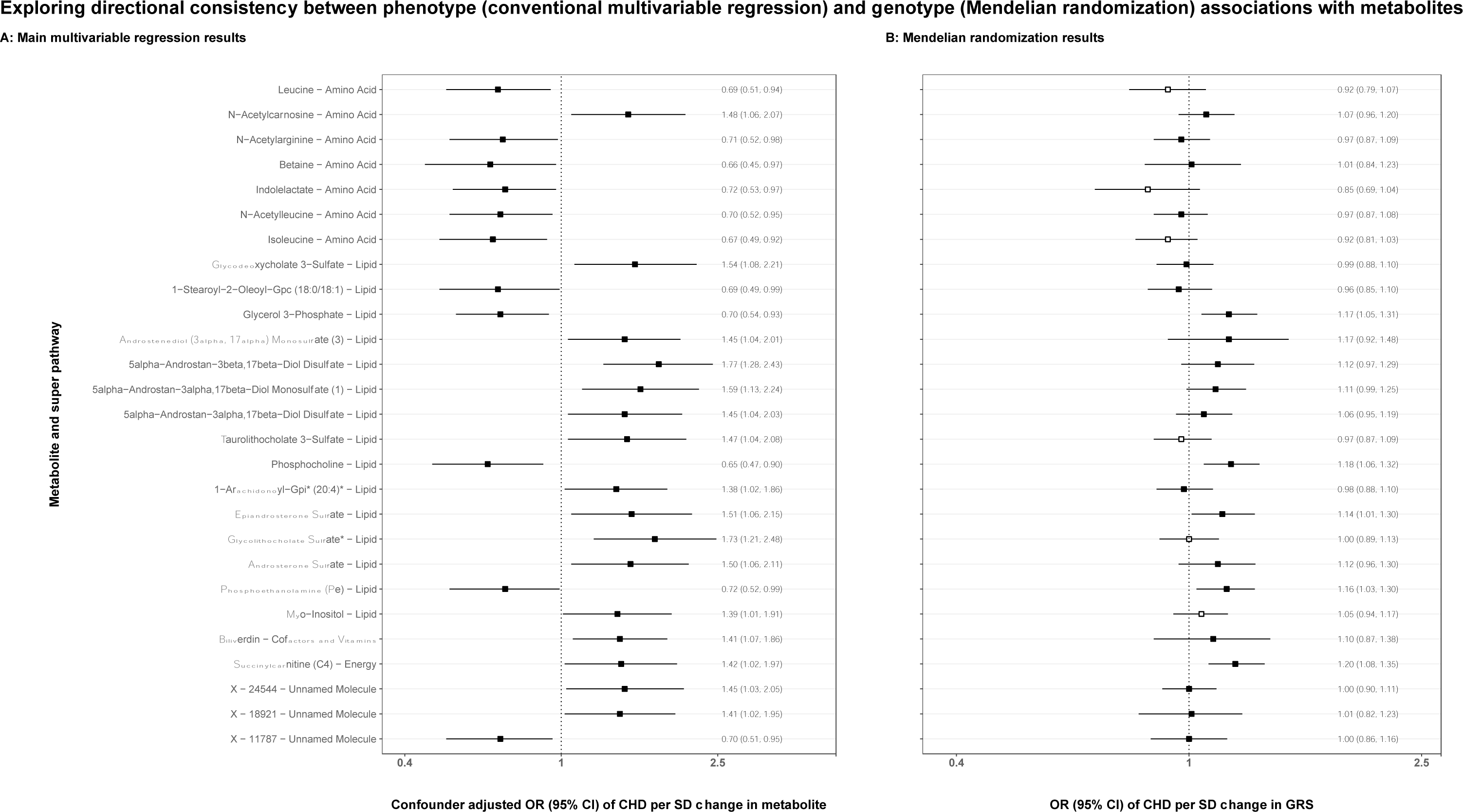
Showing results comparing the main confounder adjusted associations of maternal metabolites with offspring CHDs (Panel A: N = 2,391 & N CHD cases = 42in the Born in Bradford cohort) to the Mendelian randomization analyses of maternal genetic risk scores and offspring CHDs (Panel B: N = 38,662 & N CHD cases = 319 across 3 cohorts). N.B. results from each analysis are presented on different scales; we are not attempting to quantify estimates in the MR analyses, the aim is to compare the direction of effect. The confounder adjusted associations are as above in Figure 2. The MR analyses are adjusted for the top 10 genetic principal components and genetic batches in MoBa. In Panel B, the metabolite genetic risk scores filled with white appeared to be non-specific for the metabolite we were trying to instrument (i.e. the risk score relates to several other metabolites more strongly than the specific named metabolite).The metabolites filled in black were either metabolite-specific or specific to the metabolite and other correlated metabolites (see scatter plots in Figure S4). The results were pooled using random effects meta-analyses; individual study results and P-values for heterogeneity are shown in Supplementary Table S8. Abbreviations: BiB, Born in Bradford; CHD, congenital heart disease; GRS, genetic risk score; MR, Mendelian randomization; OR, odds ratio; CI, confidence interval.

MR analyses replicated and provided causal evidence for a potential protective effect of higher levels of the amino acids leucine, indolelactate and isoleucine on CHD, but for the other amino acids MR results were either very close to the null or in the opposite direction (**Figure 3**). Seven of the lipid-related metabolites that were positively associated in multivariable regression were also replicated in MR analyses (6 of which were highly correlated androgenic steroid metabolites), as was the energy related metabolite succinylcarnitine (**Figure 3**). For the 11 metabolites where we consider the MR GRS analyses providing some evidence of replication and a potential causal effect, 7 of the GRSs were specific for the metabolite alone and/or also for its correlates. Individual study results and P-values for heterogeneity are included in Supplementary File 2 (Table S8). MR results were largely unchanged when excluding BiB from analyses (Table S8) and when adjusting for offspring genotype (Table S9; Supplementary File 2).

## Discussion

Maternal metabolism is important for healthy fetal growth and development. To our knowledge no previous study has examined the association of detailed maternal metabolites with risk of CHD within a causal framework. In this novel study we found 44 metabolites (of 923) suggestively associated with CHD. These included metabolites related to amino acids, lipids, co-factors and vitamins, unknown molecules, xenobiotics, nucleotides and energy. In separate xenobiotics analyses, there was some evidence that metabolites related to aspirin and saccharine may increase odds of CHD, whereas metabolites related to plant food components may reduce odds. Two of the amino acids, where it was possible to explore replication, replicated in BiB in larger numbers using an alternative metabolomics platform. In MR analyses, there was directional consistency for 11/27 metabolites that could be explored with this method. We found that maternal amino acid metabolism during pregnancy, several lipids (more specifically androgenic steroids), and levels of succinylcarnitine could be important contributing factors to offspring CHD risk.

9 out of the 10 amino acids suggestively associated with CHDs were negatively associated suggesting that deficiencies in certain amino acids during pregnancy could contribute to offspring CHDs. Previous research found that amino acid concentrations measured in amniotic fluid were lower in patients with CHDs ^39^, a similar pattern to what we find here. We were able to replicate our findings for isoleucine and leucine in larger numbers in BiB which improves the confidence in the findings. The MR analyses also provided evidence to support the direction of association for these metabolites. However, the GRSs for isoleucine and leucine were non-specific and so these results should be treated with caution.

18 of the 44 maternal metabolites suggestively associated with CHD were part of the lipid super pathway, which is the most common super pathway measured by the Metabolon platform. Previous work reported that an abnormal lipid profile (defined as elevated cholesterol and apolipoprotein B) ^5^, abnormal lipid metabolism (defined as a disturbance in phosphatidyl-choline and various sphingolipids and choline metabolism) ^13^ and high maternal blood lipids ^6^ are a feature of CHD pregnancies. We were able to take forward 15 (out of 18) of the lipid metabolites and replicated the direction of effect for 7. All except 1 of these 7 replicated metabolites were androgenic steroids and so were highly correlated. Steroids are important for numerous functions during gestation, particularly for normal placental function ^40^. Here we present evidence of a potential causal effect (associated in metabolomic analyses with consistent direction of effect in MR analyses) of positive associations between maternal gestational androgenic steroid metabolites and offspring CHDs.

Levels of bilirubin and biliverdin were positively associated with CHDs. MR analyses were only possible for biliverdin but were inconclusive with wide confidence intervals. Thus, these two compounds, which are involved in heme catabolism, should be further investigated for a possible role in CHD development. Levels of succinylcarnitine were also positively associated with CHDs and we found good replications in MR analyses with consistent directions of effect and a GRS that appeared highly specific for succinylcarnitine. Succinylcarnitine is an acylcarnitine which are a group of metabolites responsible for beta oxidation of fatty acids and mitochondrial function ^41^. It is well documented that fatty acids play an important role in embryonic and fetal development ^42,43^. We included analyses of partially characterised and unknown metabolites in our results as with increasing evidence from genomic studies, previously unknown metabolites are having their function identified. With future studies identifying the function of some of these unknown/partially characterized metabolites, our results could shed light on the aetiology of CHDs.

A key strength of this study is the unique data that we have in BiB to support novel analyses of associations of a wide range of maternal metabolic paths with offspring CHD risk. We were not able to identify any other study with such data. However, we realised, even before analyses, that we would have limited statistical power with just 46 CHD cases. This motivated us to think about ways of trying to replicate any findings in larger samples either through finding measures of the same metabolites available from other assays in larger samples or using GRSs as instruments for the metabolites. In the initial multivariable regression analyses we adjusted for potential confounders. We defined suggestive associations based on a p-value threshold < 0.05, i.e., not taking account of multiple testing. When we apply a Bonferroni corrected threshold (P < 0.0001) none of the associations pass this (Supplementary Table S7; Supplementary File 2). Given the novel nature of this study and use of the initial multivariable regression in BiB to select associations for further follow-up (replication and MR), we felt this was appropriate. As with any ‘screening’ for further analyses we wanted to ensure that we would not miss potential causal effects. We recognise that selecting results based on a p-value threshold is problematic as some associations with higher p-values might have associations of a magnitude that could be clinically important, but there would also be potential for several false positives. Also, we limited MR analyses only to those metabolites that associated with p < 0.05 rather than undertaking these analyses on all of the 923 metabolites. Our reason for this was that having searched for all studies with maternal genome wide data and offspring CHD outcomes we identified only three cohorts and recognised that for MR analyses pooled results from these might also have limited power. The limited power in both multivariable and MR analyses also meant that we could only examine associations with any CHD and not subtypes.

MR analyses are sensitive to their assumptions that the GRS is statistically strongly associated with the metabolite in pregnancy. We examined associations of these with pregnancy metabolite levels and are careful in our interpretation of results in relation to this. Methods that are available for exploring potential bias due to horizontal pleiotropy in two-sample MR were not possible here. We know that many of the 923 metabolites will be biologically related to each other and with our sample size it would be difficult to robustly distinguish effects of correlated metabolites. We explored this by examining the strength of association (proportion of variation explained) of each of the 27 GRSs with all other metabolites available in BiB dataset 2. Stronger or similar associations with other metabolites would suggest that the GRS is not a specific instrument for the metabolite that we are using it for. In this case this could be because of known biological relations. For example, we know biologically that many of the lipid metabolites are related to each other, and we saw this with similar proportions of variation explained by the GRS of the androgenic steroid lipid metabolites with other androgenic steroid lipid metabolites. As such we would interpret results for these metabolites as supporting an effect of maternal androgenic steroid metabolites on CHD, but we cannot be specific about which ones are driving this. Similar or stronger variation of a GRS for other metabolites could be related to vertical pleiotropy, i.e., the metabolite for which the GRS is instrumenting strongly influences other metabolites that are related to CHD with the other metabolites partly mediating the effect of the focused metabolite. This would not bias the result. However, this could also occur with horizontal pleiotropy where the GRS, independently of the metabolite of interest, influences other metabolites that are risk factors for CHD. With our current data we are not able to distinguish between these two.

A further limitation of this study is that maternal plasma/serum metabolomics data were derived at a single timepoint around 26-28 weeks’ gestation. Fetal cardiac development starts early in pregnancy and much of the development occurs in the first trimester ^44^. Here, we are assuming that metabolite levels around 26-28 weeks’ gestation are good proxies for levels in early pregnancy - when the offspring heart is forming. Previous work has shown that between person differences throughout pregnancy remain largely consistent (i.e., those with a high level of a metabolite in early pregnancy tend to have a similarly high level of a metabolite in later pregnancy) ^45^. Similarly, and worth mentioning, the effects obtained from MR studies are often interpreted as the lifetime effect of the exposure (metabolites) in question ^46^.

In summary, we have used metabolomics data obtained during pregnancy to explore how the maternal metabolome may contribute to offspring CHDs. We found evidence that amino acid metabolism during pregnancy, several lipids (more specifically androgenic steroids), and levels of succinylcarnitine could be important contributing factors. Our analysis pipeline, which involved seeking replication of metabolite associations by harnessing large-scale GWAS data, provides scope to improve the reliability of findings and should prove to be more useful as these datasets continue to grow. Metabolomics could prove to be an important tool for identifying biological pathways that may lead to identification of prevention targets to decrease the disease burden of CHDs. To do this, future research will require international collaboration of more and larger studies with detailed metabolomics data in pregnancy, ideally with some of these having repeat measures across pregnancy and offspring CHD data.

## Data Availability

Before you contact BiB study, please make sure you have read the Guidance for Collaborators: https://borninbradford.nhs.uk/research/guidance-for-collaborators/). The ALSPAC data management plan (http://www.bristol.ac.uk/alspac/researchers/data-access/documents/alspac-data-management-plan.pdf) describes in detail the policy on data sharing, which is through a system of managed open access. Scientists are encouraged to make use of the BiB study data, which are available through a system of managed open access. Please note that the study website contains details of all the data that is available through a fully searchable data dictionary and variable search tool" and reference the following webpage: http://www.bristol.ac.uk/alspac/researchers/our-data/. MoBa data are used by researchers and research groups at both the Norwegian Institute of Public Health and other research institutions nationally and internationally. The research must adhere to the aims of MoBa and the participants' given consent. All use of data and biological material from MoBa is subject to Norwegian legislation. More information can be found on the study website (https://www.fhi.no/en/studies/moba/for-forskere-artikler/research-and-data-access/).

## Ethical approval and consent to participate

Ethical approval for ALSPAC was obtained from the ALSPAC Law and Ethics committee and local research ethics committees (NHS Haydock REC: 10/H1010/70). Informed consent for the use of data collected via questionnaires and clinics was obtained from participants following the recommendations of the ALSPAC Ethics and Law Committee at the time. At age 18, study children were sent ‘fair processing’ materials describing ALSPAC’s intended use of their health and administrative records and were given clear means to consent or object via a written form. Data were not extracted for participants who objected, or who were not sent fair processing materials. For BiB, Ethics approval has been obtained for the main platform study and all of the individual sub studies from the Bradford Research Ethics Committee. Written consent was obtained from all participants. The establishment of MoBa and initial data collection was based on a license from the Norwegian Data Protection Agency and approval from The Regional Committees for Medical and Health Research Ethics. The MoBa cohort is now based on regulations related to the Norwegian Health Registry Act.

## Availability of data and materials

The ALSPAC data management plan (http://www.bristol.ac.uk/alspac/researchers/data-access/documents/alspac-data-management-plan.pdf) describes in detail the policy on data sharing, which is through a system of managed open access. Scientists are encouraged to make use of the BiB study data, which are available through a system of managed open access. Please note that the study website contains details of all the data that is available through a fully searchable data dictionary and variable search tool” and reference the following webpage: http://www.bristol.ac.uk/alspac/researchers/our-data/. Before you contact BiB study, please make sure you have read the Guidance for Collaborators: https://borninbradford.nhs.uk/research/guidance-for-collaborators/). MoBa data are used by researchers and research groups at both the Norwegian Institute of Public Health and other research institutions nationally and internationally. The research must adhere to the aims of MoBa and the participants’ given consent. All use of data and biological material from MoBa is subject to Norwegian legislation. More information can be found on the study website (https://www.fhi.no/en/studies/moba/for-forskere-artikler/research-and-data-access/).

## Acknowledgements

BiB (Born in Bradford) study is only possible because of the enthusiasm and commitment of the children and parents. We are grateful to all the participants, practitioners, and researchers who have made BiB study happen. We are extremely grateful to all the families who took part in The Aovn Longitudinal Study of Parents and Children (ALSPAC) study, the midwives for their help in recruiting them, and the whole ALSPAC team, which includes interviewers, computer and laboratory technicians, clerical workers, research scientists, volunteers, managers, receptionists and nurses. The Norwegian Mother, Father and Child Cohort Study is supported by the Norwegian Ministry of Health and Care Services and the Ministry of Education and Research. We are grateful to all the participating families in Norway who take part in this on-going cohort study. We thank the Norwegian Institute of Public Health (NIPH) for generating high-quality genomic data. This research is part of the HARVEST collaboration, supported by the Research Council of Norway (#229624). We also thank the NORMENT Centre for providing genotype data, funded by the Research Council of Norway (#223273), South East Norway Health Authorities and Stiftelsen Kristian Gerhard Jebsen. We further thank the Center for Diabetes Research, the University of Bergen for providing genotype data and performing quality control and imputation of the data funded by the ERC AdG project SELECTionPREDISPOSED, Stiftelsen Kristian Gerhard Jebsen, Trond Mohn Foundation, the Research Council of Norway, the Novo Nordisk Foundation, the University of Bergen, and the Western Norway Health Authorities. We are grateful to all the participants who have been part of the EPIC-Norfolk metabolomics project and to the many members of the study teams at the University of Cambridge who have enabled this research.

