## Supplementary File 1 for "The relationship of maternal gestational mass spectrometry-derived metabolites with offspring congenital heart disease: results from multivariable and Mendelian randomization analyses"


- Figure S1: Metabolomics flow chart – Page 2.
- Supplementary Text and Methods – Pages 3-8.
    - Text S1, Text S2, Table S1, Text S3.
- Figure S2: Genetic analysis flow chart – Page 9.
- Supplementary results – Pages 10-21.
    - Tables S2-S4, Figures S3-S4.


Larger numerical results tables (Tables S5-S9) can be found in an Excel file (Supplementary File 2).

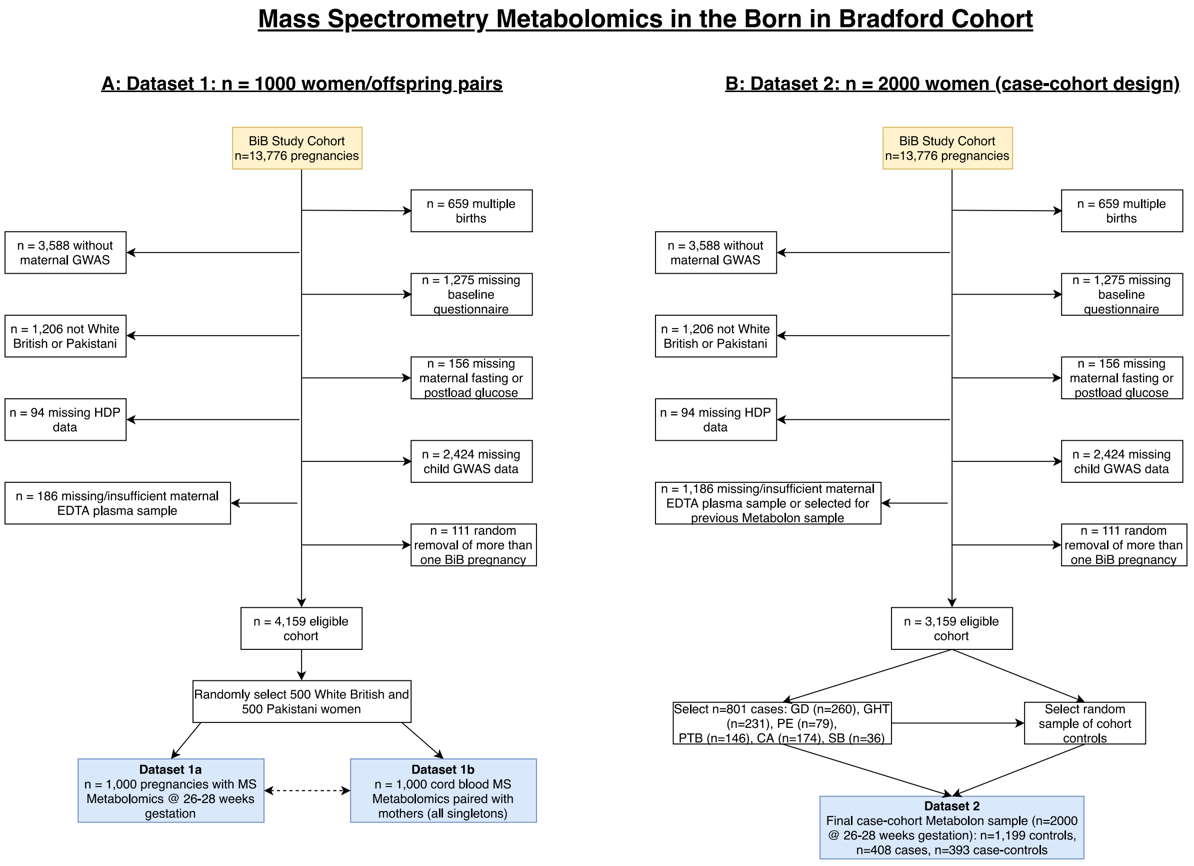


**Figure S1.** Illustrating the flow of participants into the Metabolon datasets in the Born in Bradford cohort. Panel A shows dataset 1 which includes 1,000 pregnancies and infants with MS metabolomics during pregnancy (26–28 weeks' gestation (dataset 1a)) and in cord blood (dataset 1b; dataset 1b is not included in any analyses in the present study). 1,000 women were selected on the basis that they had stored fasting plasma, a useable cord blood sample and genome wide data on both mother and offspring. Following these inclusion criteria, 500 women were selected at random from the two largest ethnic groups: White British and Pakistani, which make up 85% of the cohort. Panel B shows dataset 2 which includes 2,000 pregnancies (26–28 weeks' gestation) with MS metabolomics within a case-cohort design. This consisted of a cohort with an over-sampling of all cases of six pregnancy / perinatal outcomes. Abbreviations: MS, mass spectrometry; BiB, Born in Bradford; GWAS, genome wide association study; EDTA, ethylenediaminetetraacetic acid; HDP, hypertensive disorders of pregnancy; GD, gestational diabetes; GHT, gestational hypertension; PE, pre-eclampsia, PTB, preterm birth; CA, congenital anomaly; SB, still birth. Taken with permission from Taylor & McBride et al:

https://doi.org/10.12688/wellcomeopenres.16341.2.

**Text S1. Confounder data**

Age was obtained for all women at pregnancy booking. Ethnicity was self-reported by the mother at her recruitment questionnaire interview and based on the UK Office of National Statistics guidance. For women who did not have ethnicity data collected at the recruitment interview, data were abstracted from primary care medical records, which use a similar categorization. Women classified as South Asian included those who indicated they were Pakistani, Indian, or Bangladeshi. Women classified as White European included those who indicated that they were White British or other White European origin. Parity was categorized as having one or more previous pregnancies (multiparous (yes)) or no previous pregnancy (nulliparous (no)). We used the residential 2010 index of multiple deprivation (IMD) score presented as quintiles as a marker of socioeconomic position (SEP). Height was measured at recruitment (26–28 weeks' gestation) using a Leicester Height Measure (*Seca*, London, UK). Maternal BMI was calculated using the height measured at recruitment and weight measured at first antenatal clinic visit (approximately 12 weeks' gestation), and it was also extracted from medical records. All women were recruited from the same hospital which used *Seca* two-in-one scales (Harlow Healthcare Ltd., London, UK) to measure weight. For smoking and alcohol, women were asked the number of cigarettes smoked per day during pregnancy in the first questionnaire (26-28 weeks' gestation). We then assigned women as smokers and non-smokers. Women were asked whether they drank alcohol during pregnancy or 3 months before and assigned drinkers and non-drinkers.

**Text S2. Defining congenital heart disease**

***BiB***

In the BiB cohort, there were two separate sources to identify CAs. Both sources were used in this study: (i) CAs up to 5 years of age, identified in GP records by Bishop et al [30] following EUROCAT guidelines. ICD-10 codes were mapped to clinical term (CT)-V3 codes prior to extraction from GP records. (ii) Data extracted from the Yorkshire and Humber CAs register database. Data were ICD-10 coded. All of these were confirmed postnatally. BiB includes data on the birth outcome of each child (live birth, miscarriage, still birth). Therefore, diagnoses were not necessarily restricted to live born children. However, there is the possibility that some women would have terminated the pregnancy after the 12- or 20-week scans which would lead to an under-representation of congenital anomaly cases.

***ALSPAC***

Case ascertainment of CAs in the ALSPAC cohort has been described in detail in a recently published data note [29]. Data were combined from multiple sources: NHS records (primary care, paediatric cardiology database, data on fetal deaths and local child health services), midwifery and birth records and maternal self-report via child-based questionnaires. Each source was coded using ICD-10 codes. By combining sources, there would be a greater possibility of capturing all of possible cases within the cohort. The majority of cases of CAs were identified by primary care records (79% for any CA and 68% for any CHD). We included diagnoses made at any age (from birth up until age 25/26). There were no restrictions in cases of CAs in ALSPAC, we included all cases whether live-born or not. However, it is possible that some cases that were terminated earlier in pregnancy were missed due to them never having an NHS number and thus not being identified through record linkage.

***MoBa***

Information on whether a child had a CHD or not was obtained though linkage to the Medical Birth Registry of Norway (MBRN). All maternity units in Norway must notify births to the MBRN. Further information can be found in the publication by Leirgul et al (https://doi.org/10.1016/j.ahj.2014.07.030). The notification form includes the name and personal identity number of the child and parents, as well as information about maternal health before and during pregnancy, and any complications during pregnancy or at birth, including the presence of any heart defects. The MBRN contains information on all births and pregnancies ended after the 12th week of gestation, including stillbirths and abortions after the 12th week, including on heart defects. Heart defects are registered in the MBRN through notifications from clinical staff identifying these defects at delivery or any hospital in patient treatments occurring immediately after birth until the child is discharged. The medical notification is made at discharge, which can be several months after birth. Details of the notified heart defects, such as specific diagnosis or treatment are not provided. Whilst most of the heart defects would have been diagnosed at birth it is possible that some children were admitted to hospital after delivery for non-specific reasons of for diagnoses that at the time were not considered to be related to a heart defect. Therefore, we considered MoBa diagnoses to have been made between birth and 6 months (few would remain in hospital after this length).

**Table S1.** Subcategories of CHD.

| Category | CHDs included/excl | ICD-10 codes |
|---|---|---|
| All CHDs | Any CHD as defined by EUROCAT*<br>Patent ductus arteriosus with gestational age < 37 weeks not considered a CHD case.<br>Peripheral pulmonary artery stenosis with gestational age < 37weeks not considered as a CHD case. | Q20-Q25, Q260, Q262-Q269** |

* Definitions taken from here: https://eu-rd-platform.jrc.ec.europa.eu/sites/default/files/EUROCAT-Guide-1.4-Section-3.3.pdf
**Q250 and Q256 not a case if isolated and GA<37weeks
Abbreviations: CHD, congenital heart disease; ICD, international classification of disease; EUROCAT, European surveillance of congenital anomalies.

**Text S3. Genetic data methods**

***Born in Bradford (BiB)***

The samples of the BiB cohort (mothers and offspring) were processed on three different type of Illumina chips: HumanCoreExome12v1.0, HumanCoreExome12v1.1 and HumanCoreExome24v1.0. The pre-processing of samples was done separately for the three chips. Problematic samples which had a Call Rate < 0.95 were removed. Poorly performing SNPs determined by a set of quality matrices were zeroed.

*BiB Illumina HumanCoreExome: PLINK and filtering*

GenomeStudio output files were converted to PLINK format and subsequently filtered. SNPs where >=20% of individuals were missing genotype were removed. Individuals with >=10% missing genotypes were removed. A further pass over genotype rate was performed, removing SNPs where over 20% are missing genotype. Following inspection of plink *--missing output*, individuals with > 1% missing genotypes were removed. The final pass over genotype rate, removed SNPs where over 0.5% were missing genotype. The final pass over missingness per individual, removed individuals with over 0.5% missing genotypes.

*Quality control and imputation*

From each of the 3 genotyping sets any individual or SNP missing >3% of their data was dropped and the datasets combined. Genetic duplicates were removed. Reported first degree relatives (mother-child, father-child, child-child siblings) were checked to see if they looked genetically like first degree relatives. If there was no such evidence of this, they were removed. Mother-child discrepancies between phenotype and genotype were removed. People who looked genetically to be clearly South Asian or White British from the principal component analysis (PCA) but had a different ethnicity phenotype were removed. Based on a combination of PCA and reported ethnicity there were two subsets of individuals – white European and south Asian. As a sensitivity analysis, all of the genetic analyses in BiB are repeated after stratifying by these two ethnic groups. SNPs with a minor allele frequency (MAF) <1%, genotyping rate <5% or with a deviation from Hardy–Weinberg disequilibrium (pP << 1×10−6) were removed from the analysis. Imputation was performed for Europeans and South Asians separately, both using the HRC r1.1 as the reference panel. The genotype data was uploaded to the Michigan Imputation Server to perform genotype imputation using Minimac4. Phasing was performed using Eagle v2.4. After imputation, the VCF files were downloaded from the server and BCFtools was used to remove SNPs that were not accurately imputed. Mimimac4 generates a metric (imputation accuracy $R^{\text{-squared}}$) for each variant, and variants with estimated imputation accuracy $R^2$ <0.3 were removed.

***The Avon Longitudinal Study of Parents and Children (ALSPAC)***

Mothers were genotyped on Illumina HumanHap660W quad-chip platform by Centre National de Génotypage (Évry, FR). Offspring were genotyped on Illumina HumanHap550 quad-chip platforms by the Wellcome Trust Sanger Institute (Cambridge, UK) and by the Laboratory Corporation of America (Burlington, USA) using support from 23andMe. Standard quality control was applied to SNPs and individuals. Individuals were excluded based on genotype rate (<5%), sex mismatch, high heterozygosity and cryptic relatedness [defined as identity-by-descent (IBD) >0.125]. In order to remove individuals of non-European descent, principal components (PCs) were derived from linkage disequilibrium-pruned SNPs with MAF >0.01 using plink. Individuals laying 5 standard deviations beyond the 1000 Genomes European population PCs 1 and 2 centroid were excluded. SNPs with a minor allele frequency (MAF) <1%, genotyping rate <5% or with a deviation from Hardy–Weinberg disequilibrium (pP << 1×10−6) were removed from the analysis. Using this QC'd dataset, a list of unrelated mothers was created using an IBD cut-off of 0.05. For imputation, genotypes of ALSPAC mothers and children were combined. Haplotypes were estimated using ShapeIT (v2. r644), which utilises relatedness during phasing. A phased version of the 1000 genomes reference panel (Phase 1, Version 3) was obtained from the Impute2 reference data repository. Imputation was performed using Impute V2.2.2 against the reference panel (all polymorphic SNPs excluding singletons), using all 2186 reference haplotypes (including non-Europeans).

***The Norwegian Mother, Father and Child Cohort (MoBa)***

Compared to other large biobanks like the UK Biobank, where considerable funding was secured upfront allowing for genotyping their entire cohort in a single effort, genotyping in MoBa have had to rely on several projects - each contributing with resources to genotype subsets of MoBa over the last decade. Consequently, genotyping was performed years apart at different labs using different arrays. We used data from MoBaGenetics 1.0. There is an openly available comprehensive GitHub page that documents all quality control for all releases of genetic data in the MoBa cohort (https://github.com/folkehelseinstituttet/mobagen). In this study we used data from the following batches: NORMENT, ROTTERDAM, TED and HARVEST (initial N = ~98,000). 33,047 individuals were genotyped in the NORMENT sample at deCODE genetics, Reykjavik Iceland (Illumina HumanOmniExpress-24v1.0, Illumina InfiniumOmniExpress-24v1.2, & Illumina Global Screening Array MD v.1.0 + 50k custom OmniExpress overlap content array), 26,680 were genotyped in the ROTTERDAM sample at ERASMUS MC,

Rotterdam, Netherlands (Illumina Global Screening Array MD v.1.0 array), 5215 were sampled in the TED samples at deCODE genetics (Illumina InfiniumOmniExpress-24v1.2), and 32,886 were sampled in the HARVEST sample at Genomics Core Facility, Trondheim, Norway (llumina HumanCoreExome12v1.1 & Illumina HumanCoreExome24v1.0). Below, we describe the methods and QC for the merged dataset used in the present study.

Quality control (QC) and imputation was performed to align with current best-practice QC protocols in human genetics and the family-based pipeline Picopili. The primary software used for the QC was PLINK 1.9 and KING 2.2.5. To identify core subpopulations filtering of was performed for minor allele frequency of 1%, SNP and individual call rate of 95%, and Hardy-Weinberg Equilibrium (HWE) p-value of 0.001. Principle component (PC) analysis with 1000 Genomes phase 1 data was used to identify the European, Asian, and African core subpopulations.

Pre-imputation QC was performed for each of the core subpopulations on the SNP and individual level. QC on a SNP level involved filtering for 0.5% MAF, 95% call rate, HWE p-value 0.000001, discordant in duplicate pairs, association with genotype plate and genotype batch at p-value 0.001. Individual level QC was performed by filtering for heterozygosity outliers Fhet ± 0.2, erroneous sex assignment, known relatedness, cryptic relatedness, identity-by-decent (PI_HAT threshold of 0.15), and PC outliers both with and without 1000 Genomes. Mendel errors were assessed for families with a minimum of one PO duo. Families with more than 5% Mendel errors and SNPs with more than 1% of Mendel errors were removed, while other minor Mendel errors were zeroed out. Batches that were genotyped using the same array were merged (keeping only SNPs present in all batches) and the pre-imputation QC was performed on the merged batches.

Phasing and imputation was performed using the publicly available Haplotype Reference Consortium data. Phasing was performed using SHAPEIT2 with the duoHMM algorithm to incorporate the pedigree information into the haplotype estimates. IMPUTE 4 was then used to perform imputation. Dosage data was then converted to best-guess, hard call genotype data with an imputation quality score (INFO) of 0.8 and default PLINK certainty of 0.9. Post-imputation QC was then performed following the steps outlined in the pre-imputation QC. To ensure the across batch relatedness (both known, such as PO and FS relationships, and unknown, such as sibships within the parent generation) was accounted for in all analyses the three imputation batches were merged, and post-imputation QC was performed on the overall merged dataset. We removed related individuals (cryptic relatedness: IBD >0.05).

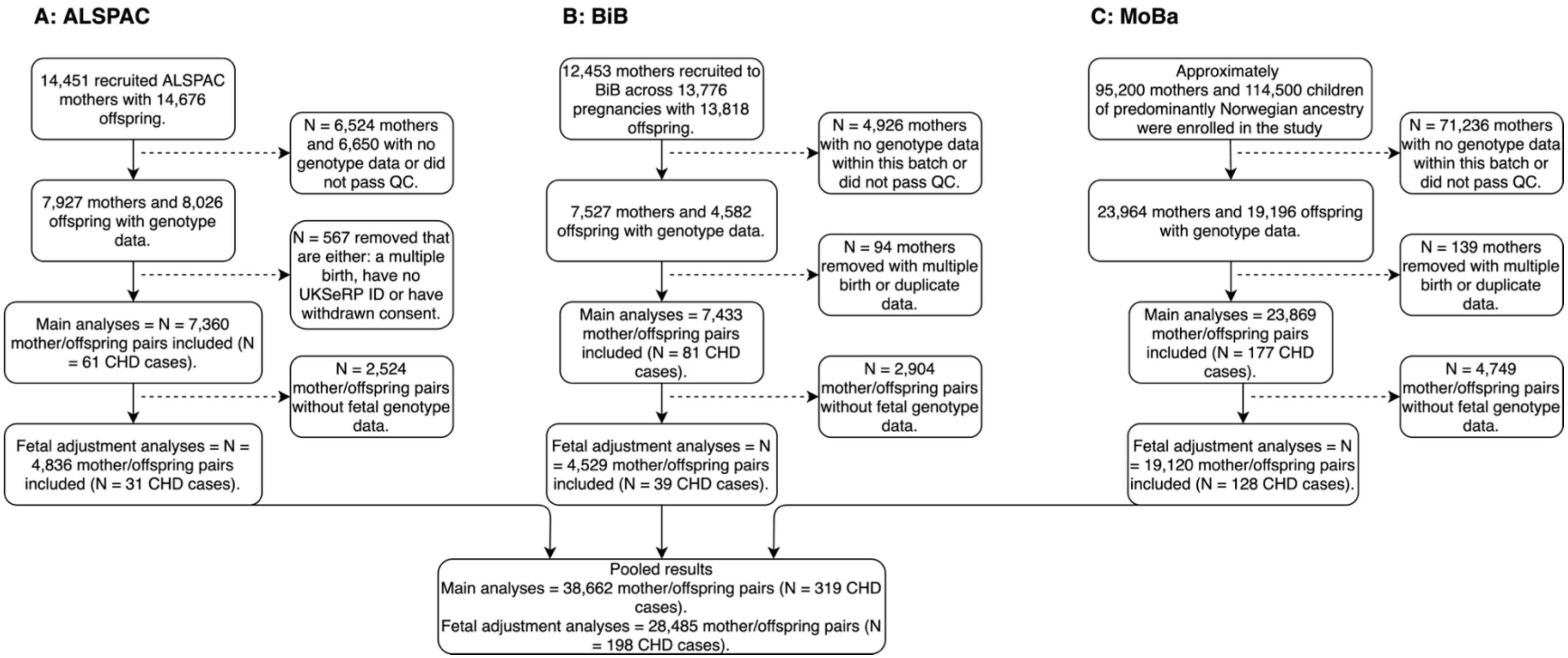


**Figure S2. An overview of included cohorts and selection of study participants for Mendelian randomization analyses.** Abbreviations: ALSPAC, Avon Longitudinal Study of Parents and Children; BiB, Born in Bradford; MoBa, Norwegian Mother, Father and Child Cohort; QC, quality control; UKSeRP, the secure research platform containing CHD data for ALSPAC; CHD, congenital heart disease; GWAS, genome-wise association study association study.

### Supplementary Results

**Table S2: Logistic regression results from the presence/absence xenobiotic analysis.**

| Metabolite | Sub pathway | $OR_c$ (95% CI) | $OR_a$ (95% CI) |
|---|---|---|---|
| saccharin | Food Component/Plant | 1.82 (0.91, 4.03) | 2.16 (1.02, 5.13) |
| salicyluric glucuronide | Drug - Analgesics, Anesthetics | 2.01 (1.05, 3.70) | 2.27 (1.16, 4.29) |
| alliin | Food Component/Plant | 0.60 (0.33, 1.10) | 0.35 (0.17, 0.73) |
| ferulic acid 4-sulfate | Food Component/Plant | 0.74 (0.40, 1.34) | 0.50 (0.25, 0.96) |
| naringenin 7-glucuronide | Food Component/Plant | 0.28 (0.05, 0.93) | 0.14 (0.01, 0.66) |
| glucuronide of piperine metabolite C17H21NO3 (5) | Food Component/Plant | 0.42 (0.20, 0.81) | 0.47 (0.22, 0.93) |
| Odds ratios are given for the presence of the metabolite during pregnancy and odds of having CHD in the offspring.<br>$OR_c$ = unadjusted odds ratio (N = 2,605 [46 CHD cases]); $OR_a$ = adjusted odds ratio (N = 2,426 [42 CHD cases]), adjusted for: maternal age, ethnicity, parity, SEP, BMI, smoking). | | | |

**Table S3. Participant characteristics for the 3 studies included in Mendelian randomization analyses.**

| Characteristic | Category | ALSPAC (N = 7,360) | BiB (N = 7,433) | MoBa (N = 23,869) |
|---|---|---|---|---|
| ***Offspring*** | | | | |
| CHD | Yes | 61 (0.8) | 81 (1.1) | 177 (0.7) |
| Sex | Male | 3,703 (50.3) | 3,818 (51.4) | 12,139 (50.9) |
| | Female | 3,657 (49.7) | 3,615 (48.6) | 11,704 (49.0) |
| ***Maternal*** | | | | |
| Age, years | | 29.2 (4.6) | 27.4 (5.6) | 30.1 (4.5) |
| Parity | Primiparous | 3,257 (46.6) | 2,963 (40.1) | 11,288 (47.3) |
| BMI, $kg/m^2$ | | 22.5 (4.2) | 26.2 (5.7) | 24.1 (4.3) |
| Ethnicity | White European | 7,360 (100.0) [a] | 3,084 (42.6) | 23,869 (100.0) [b] |
| | South Asian | - | 3,503 (48.4) | - |
| | Other | - | 656 (9.1) | - |
| Any smoking during pregnancy | Yes | 1,679 (26.1) | 1,175 (18.1) | 1,814 (8.6) |
| Any alcohol during pregnancy | Yes | 4,866 (79.9) | 1,040 (49.3) | 6,209 (31.5) |

Data are means ± SD or n (%) unless stated. % are based on data available (data were not complete).

[a] All non-white European women with ethnicity data were not included in the analysis.

[b] Individuals of non-European ancestries were removed based on principal component analysis

Abbreviations: BiB, Born in Bradford; ALSPAC, Avon Longitudinal Study of Parents and Children; MoBa, Norwegian Mother, Father and Child Cohort Study; CHD, congenital heart disease; BMI, body mass index; kg, kilograms; m, meters.

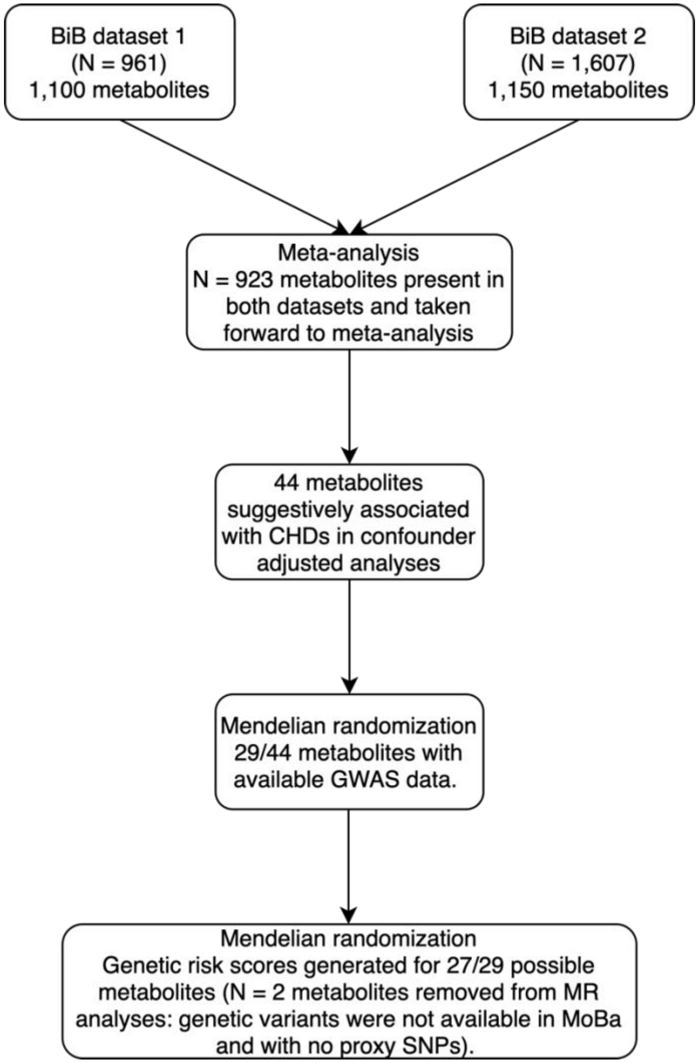


**Figure S3.** Flow chart to illustrate the analysis pipeline and selection of metabolites for Mendelian randomization. The two metabolites (Glycerophosphoethanolamine and X-24295) were the two metabolites removed from MR analyses because they had a small number of SNPs and these were not present in MoBa and had no proxy's.

**Table S4. Characteristics of maternal GRS and associations with the corresponding metabolite in BiB Dataset 2 (N = 1,326).**

| Metabolite | Super pathway | HMDB | N SNPs in GRS[a] | Coefficient (95% CI)[b] | P-Value | $R^2$ (%) | F-statistic |
|---|---|---|---|---|---|---|---|
| Isoleucine | Amino Acid | HMDB00172 | 5 | 0.06 (0.0006, 0.11) | 0.05 | 0.3 | 3.9 |
| N-Acetylleucine | Amino Acid | HMDB11756 | 2 | 0.37 (0.32, 0.43) | 9.38E-41 | 12.6 | 191.4 |
| Indolelactate | Amino Acid | HMDB00671 | 1 | 0.04 (-0.01, 0.09) | 0.15 | 0.2 | 2.0 |
| Betaine | Amino Acid | HMDB00043 | 9 | 0.19 (0.14, 0.25) | 1.23E-11 | 3.4 | 46.7 |
| N-Acetylarginine | Amino Acid | HMDB04620 | 10 | 0.59 (0.55, 0.63) | 8.55E-122 | 34.0 | 683.3 |
| N-Acetylcarnosine | Amino Acid | HMDB12881 | 9 | 0.25 (0.20, 0.30) | 1.57E-20 | 6.3 | 89.2 |
| Leucine | Amino Acid | HMDB00687 | 6 | 0.02 (-0.04, 0.07) | 0.55 | 0.3 | 0.4 |
| Myo-Inositol | Lipid | HMDB00211 | 2 | 0.02 (-0.04, 0.08) | 0.56 | 0.03 | 0.3 |
| Phosphoethanolamine (Pe) | Lipid | HMDB00224 | 2 | 0.07 (0.02, 0.12) | 0.01 | 0.5 | 6.4 |
| Androsterone Sulfate | Lipid | HMDB02759 | 11 | 0.30 (0.25, 0.36) | 1.12E-28 | 8.9 | 129.4 |
| Glycolithocholate Sulfate* | Lipid | HMDB02639 | 2 | 0.10 (0.05, 0.16) | 0.0004 | 1.0 | 12.7 |
| Epiandrosterone Sulfate | Lipid |  | 10 | 0.25 (0.20, 0.31) | 1.69E-20 | 6.3 | 89.0 |
| 1-Arachidonoyl-Gpi* (20:4)* | Lipid | HMDB61690 | 2 | 0.15 (0.10, 0.21) | 2.27E-08 | 2.3 | 31.6 |
| Phosphocholine | Lipid | HMDB01565 | 6 | 0.15 (0.10, 0.20) | 1.98E-09 | 2.7 | 36.5 |
| Taurolithocholate 3-Sulfate | Lipid | HMDB02580 | 4 | 0.10 (0.04, 0.15) | 0.0004 | 0.9 | 12.5 |
| 5alpha-Androstan-3alpha,17beta-Diol Disulfate | Lipid | - | 6 | 0.07 (0.02, 0.13) | 0.009 | 0.5 | 6.9 |
| 5alpha-Androstan-3alpha,17beta-Diol Monosulfate (1) | Lipid | - | 14 | 0.23 (0.18, 0.29) | 4.41E-17 | 5.2 | 72.5 |
| 5alpha-Sndrostan-3beta,17beta-Diol Disulfate | Lipid | HMDB00493 | 11 | 0.24 (0.19, 0.30) | 6.05E-19 | 5.8 | 81.5 |
| Androstenediol (3alpha, 17alpha) Monosulfate (3) | Lipid | - | 13 | 0.31 (0.26, 0.37) | 3.97E-30 | 9.4 | 136.7 |
| Glycerol 3-Phosphate | Lipid | HMDB00126 | 3 | 0.12 (0.07, 0.17) | 4.80E-06 | 1.6 | 21.1 |
| 1-Stearoyl-2-Oleoyl-GPC (18:0/18:1) | Lipid | HMDB08038 | 3 | 0.13 (0.08, 0.18) | 1.16E-06 | 1.8 | 23.9 |
| Glycodeoxycholate 3-Sulfate | Lipid | - | 5 | 0.20 (0.15, 0.25) | 2.18E-14 | 4.3 | 59.7 |
| Biliverdin | Cofactors and Vitamins | HMDB01008 | 3 | 0.52 (0.48, 0.57) | 3.01E-93 | 27.2 | 493.9 |
| Succinylcarnitine (C4) | Energy | HMDB61717 | 7 | 0.35 (0.30, 0.40) | 1.56E-45 | 14.1 | 216.7 |
| X - 11787 | NA | - | 5 | 0.28 (0.23, 0.34) | 3.93E-26 | 8.1 | 116.7 |
| X - 18921 | NA | - | 3 | 0.27 (0.22, 0.33) | 2.80E-25 | 7.8 | 112.5 |
| X - 24544 | NA | - | 5 | 0.19 (0.14, 0.24) | 2.03E-13 | 4.0 | 55.1 |

[a] SNPs for isoleucine and leucine taken from: https://www.nature.com/articles/s41588-020-00751-5; remainder of SNPs taken from a recent GWAS of metabolon metabolites (unpublished).
[b] Estimates from linear regression interpreted as difference in metabolite (scaled/imputed metabolites were log-transformed and presented in SD units as per manuscript methods) per SD increase in genetic risk score. MS-derived metabolomics measured using plasma taken during pregnancy around 26-28 weeks' gestation in the BiB cohort (see methods).
N = 1,326 is the number of women in BiB dataset 2 with Metabolon metabolomics data and GWAS data.
Abbreviations: HMDB, The Human Metabolome Database; SNP, single nucleotide polymorphism; GRS, genetic risk score; CI, confidence interval.

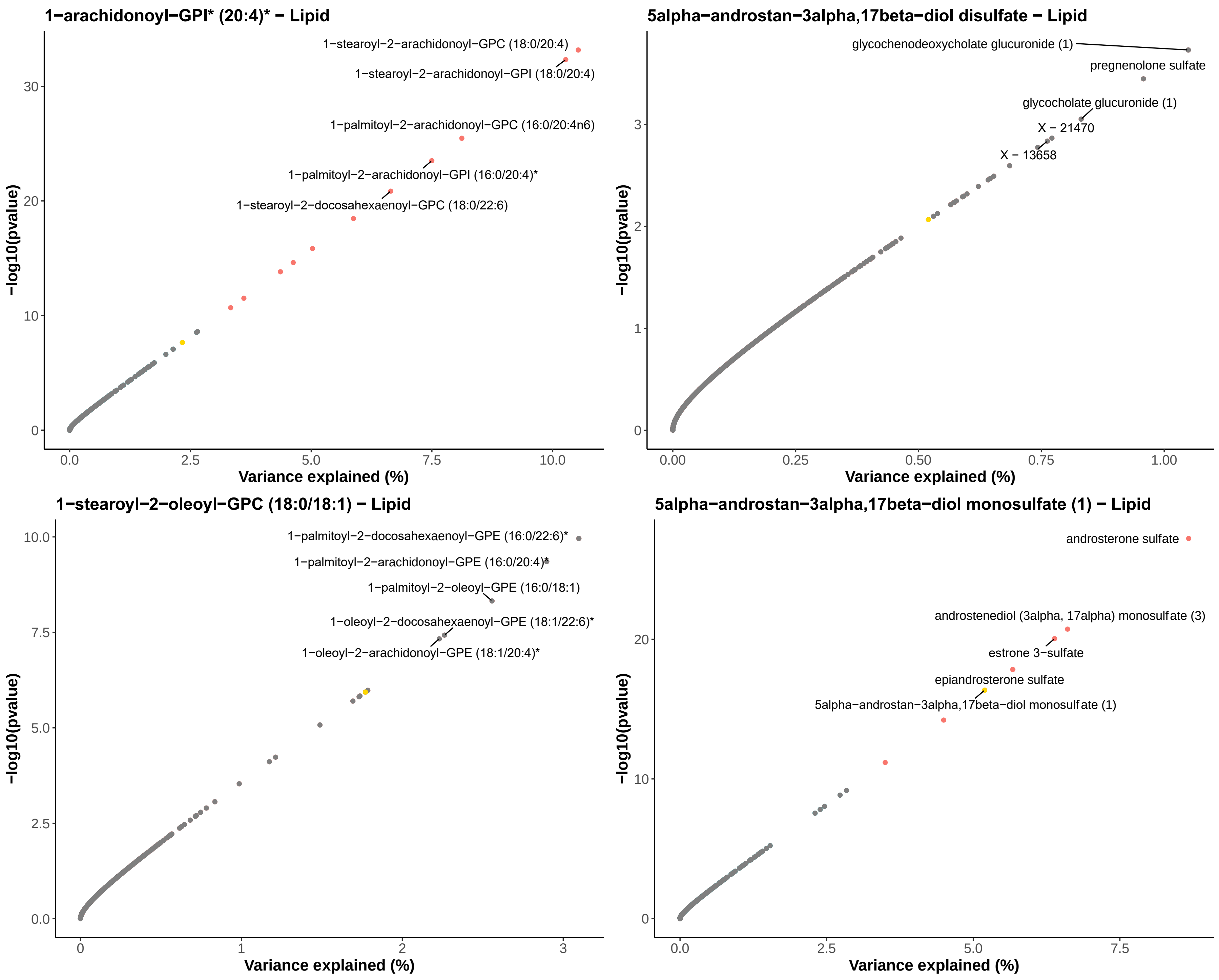

1−arachidonoyl−GPI* (20:4)* − Lipid
1−stearoyl−2−arachidonoyl−GPC (18:0/20:4)
1−stearoyl−2−arachidonoyl−GPI (18:0/20:4)
1−palmitoyl−2−arachidonoyl−GPC (16:0/20:4n6)
1−palmitoyl−2−arachidonoyl−GPI (16:0/20:4)*
1−stearoyl−2−docosahexaenoyl−GPC (18:0/22:6)
−log10(pvalue)
Variance explained (%)
5alpha−androstan−3alpha,17beta−diol disulfate − Lipid
glycochenodeoxycholate glucuronide (1)
pregnenolone sulfate
glycocholate glucuronide (1)
X − 21470
X − 13658
−log10(pvalue)
Variance explained (%)
1−stearoyl−2−oleoyl−GPC (18:0/18:1) − Lipid
1−palmitoyl−2−docosahexaenoyl−GPE (16:0/22:6)*
1−palmitoyl−2−arachidonoyl−GPE (16:0/20:4)*
1−palmitoyl−2−oleoyl−GPE (16:0/18:1)
1−oleoyl−2−docosahexaenoyl−GPE (18:1/22:6)*
1−oleoyl−2−arachidonoyl−GPE (18:1/20:4)*
−log10(pvalue)
Variance explained (%)
5alpha−androstan−3alpha,17beta−diol monosulfate (1) − Lipid
androsterone sulfate
androstenediol (3alpha, 17alpha) monosulfate (3)
estrone 3−sulfate
epiandrosterone sulfate
5alpha−androstan−3alpha,17beta−diol monosulfate (1)
−log10(pvalue)
Variance explained (%)

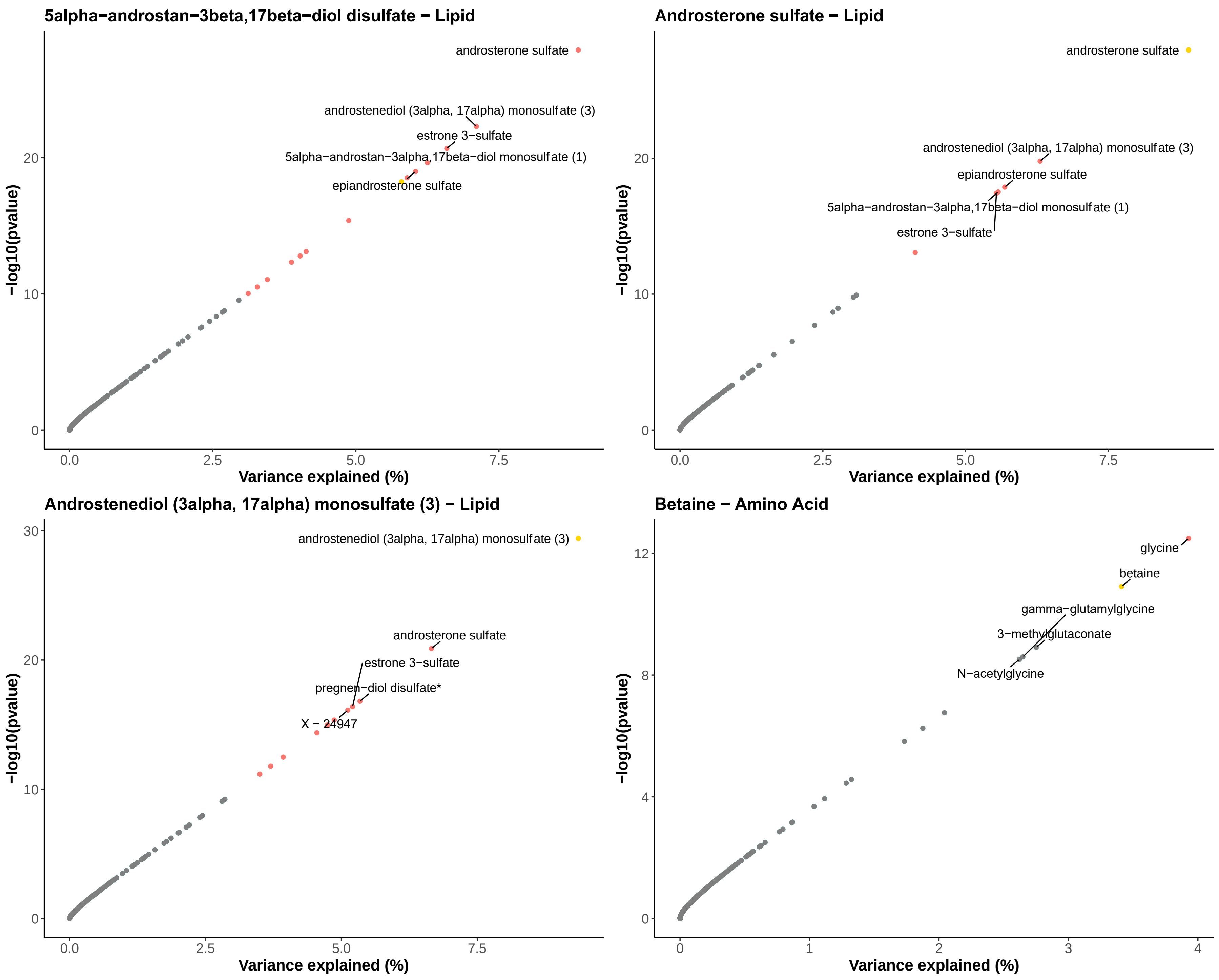
5alpha−androstan−3beta,17beta−diol disulfate − Lipid
androsterone sulfate
androstenediol (3alpha, 17alpha) monosulfate (3)
estrone 3−sulfate
5alpha−androstan−3alpha,17beta−diol monosulfate (1)
epiandrosterone sulfate
−log10(pvalue)
Variance explained (%)
0
10
20
0.0
2.5
5.0
7.5
Androsterone sulfate − Lipid
androsterone sulfate
androstenediol (3alpha, 17alpha) monosulfate (3)
epiandrosterone sulfate
5alpha−androstan−3alpha,17beta−diol monosulfate (1)
estrone 3−sulfate
−log10(pvalue)
Variance explained (%)
0
10
20
0.0
2.5
5.0
7.5
Androstenediol (3alpha, 17alpha) monosulfate (3) − Lipid
androstenediol (3alpha, 17alpha) monosulfate (3)
androsterone sulfate
estrone 3−sulfate
pregnen−diol disulfate*
X − 24947
−log10(pvalue)
Variance explained (%)
0
10
20
30
0.0
2.5
5.0
7.5
Betaine − Amino Acid
glycine
betaine
gamma−glutamylglycine
3−methylglutaconate
N−acetylglycine
−log10(pvalue)
Variance explained (%)
0
4
8
12
0
1
2
3
4

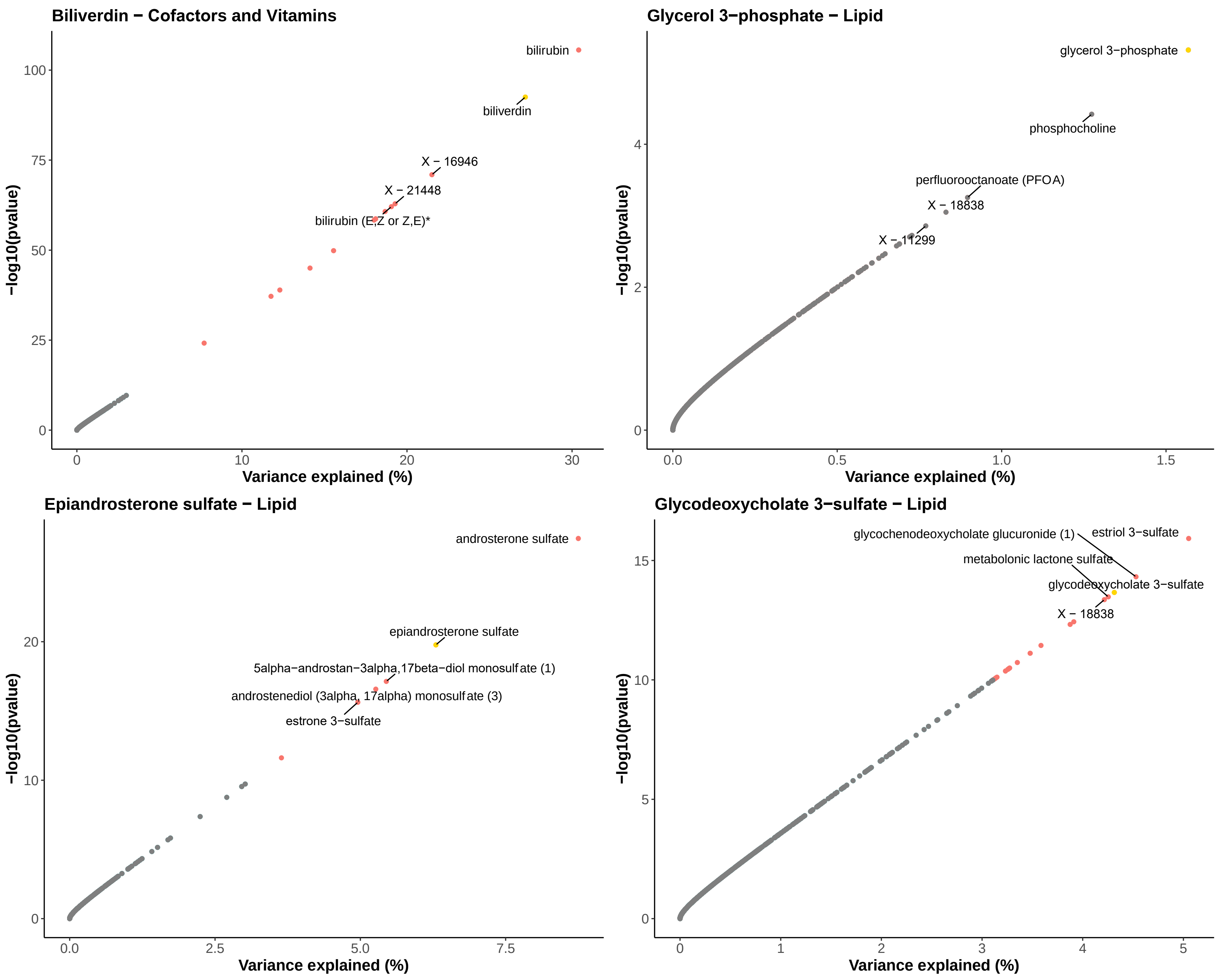
Biliverdin − Cofactors and Vitamins
bilirubin
biliverdin
X − 16946
X − 21448
bilirubin (E,Z or Z,E)*
−log10(pvalue)
Variance explained (%)
Glycerol 3−phosphate − Lipid
glycerol 3−phosphate
phosphocholine
perfluorooctanoate (PFOA)
X − 18838
X − 11299
−log10(pvalue)
Variance explained (%)
Epiandrosterone sulfate − Lipid
androsterone sulfate
epiandrosterone sulfate
5alpha−androstan−3alpha,17beta−diol monosulfate (1)
androstenediol (3alpha, 17alpha) monosulfate (3)
estrone 3−sulfate
−log10(pvalue)
Variance explained (%)
Glycodeoxycholate 3−sulfate − Lipid
glycochenodeoxycholate glucuronide (1)
estriol 3−sulfate
metabolonic lactone sulfate
glycodeoxycholate 3−sulfate
X − 18838
−log10(pvalue)
Variance explained (%)

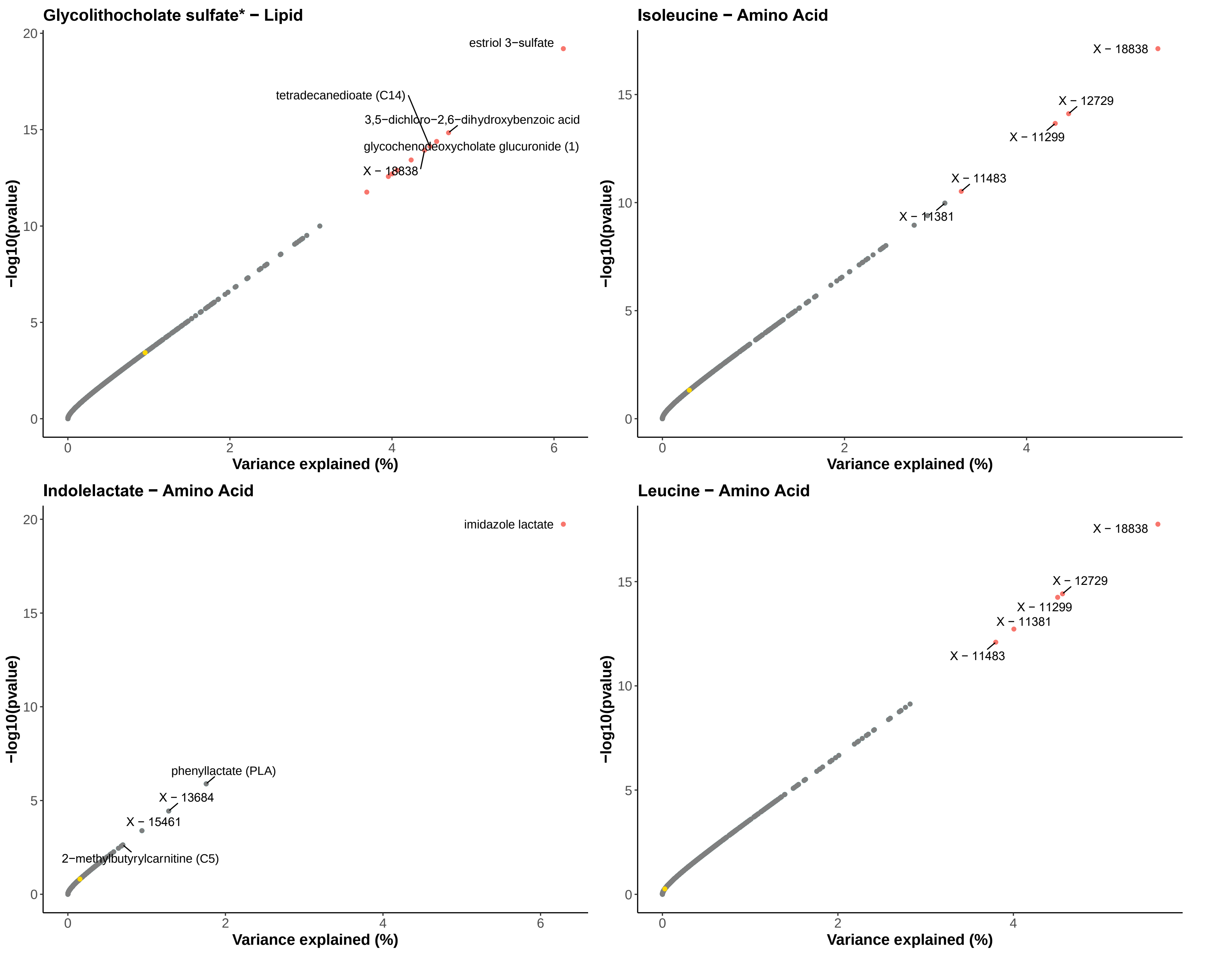
Glycolithocholate sulfate* − Lipid
estriol 3−sulfate
tetradecanedioate (C14)
3,5−dichloro−2,6−dihydroxybenzoic acid
glycochenodeoxycholate glucuronide (1)
X − 18838
−log10(pvalue)
Variance explained (%)
Isoleucine − Amino Acid
X − 18838
X − 12729
X − 11299
X − 11483
X − 11381
−log10(pvalue)
Variance explained (%)
Indolelactate − Amino Acid
imidazole lactate
phenyllactate (PLA)
X − 13684
X − 15461
2−methylbutyrylcarnitine (C5)
−log10(pvalue)
Variance explained (%)
Leucine − Amino Acid
X − 18838
X − 12729
X − 11299
X − 11381
X − 11483
−log10(pvalue)
Variance explained (%)

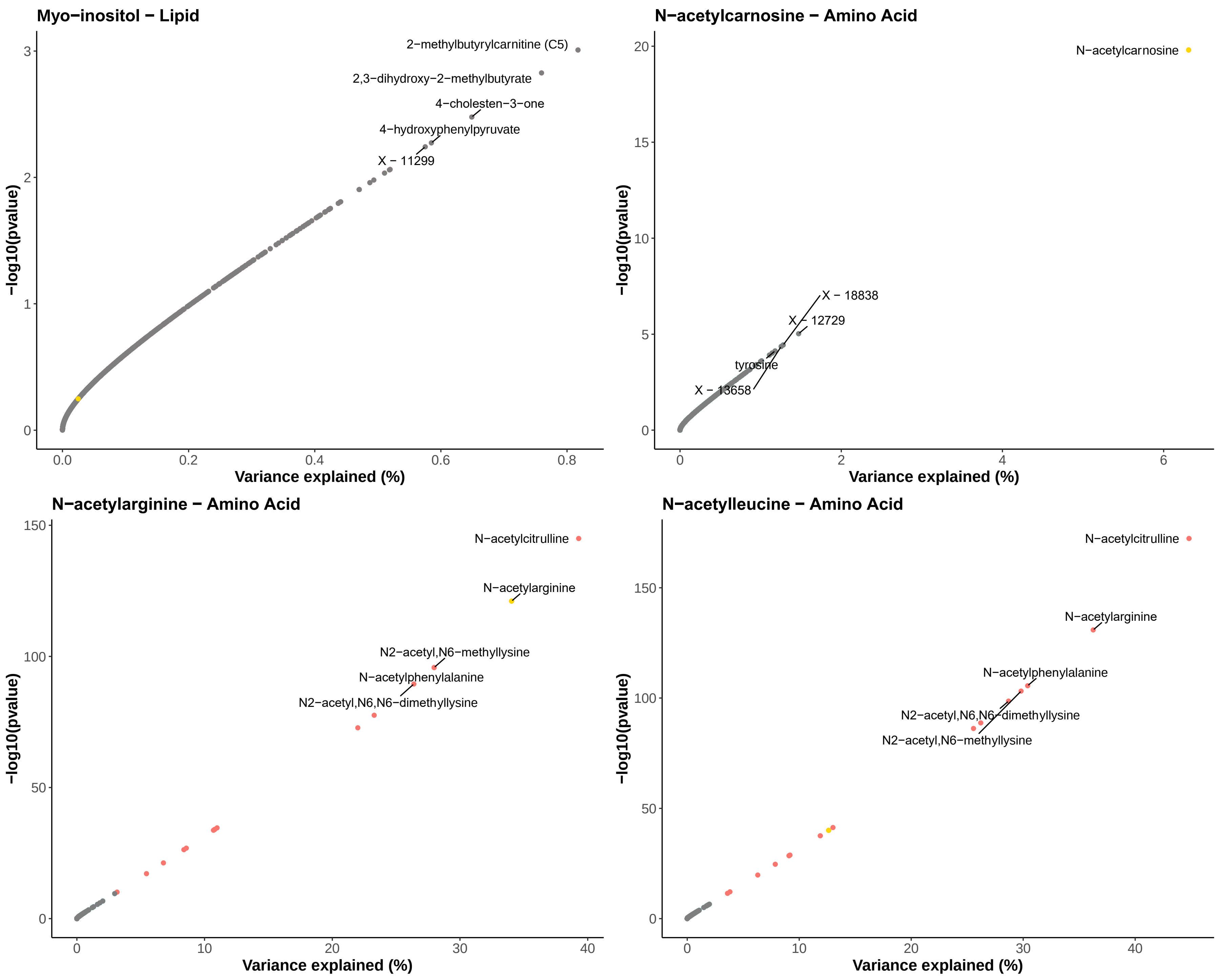

Myo−inositol − Lipid
2−methylbutyrylcarnitine (C5)
2,3−dihydroxy−2−methylbutyrate
4−cholesten−3−one
4−hydroxyphenylpyruvate
X − 11299
−log10(pvalue)
Variance explained (%)
N−acetylcarnosine − Amino Acid
N−acetylcarnosine
X − 18838
X − 12729
tyrosine
X − 13658
−log10(pvalue)
Variance explained (%)
N−acetylarginine − Amino Acid
N−acetylcitrulline
N−acetylarginine
N2−acetyl,N6−methyllysine
N−acetylphenylalanine
N2−acetyl,N6,N6−dimethyllysine
−log10(pvalue)
Variance explained (%)
N−acetylleucine − Amino Acid
N−acetylcitrulline
N−acetylarginine
N−acetylphenylalanine
N2−acetyl,N6,N6−dimethyllysine
N2−acetyl,N6−methyllysine
−log10(pvalue)
Variance explained (%)

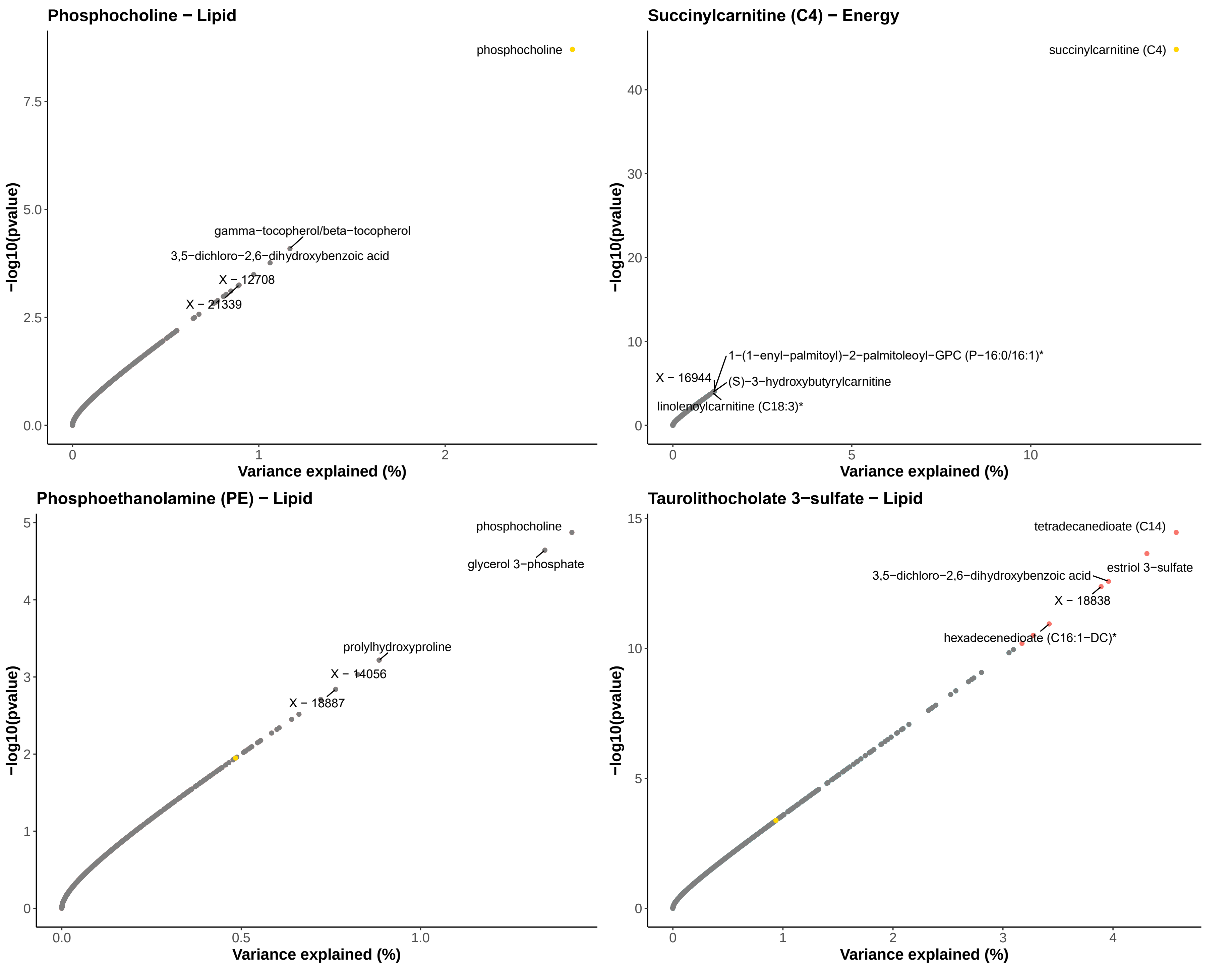
Phosphocholine − Lipid
phosphocholine
gamma−tocopherol/beta−tocopherol
3,5−dichloro−2,6−dihydroxybenzoic acid
X − 12708
X − 21339
−log10(pvalue)
Variance explained (%)
Succinylcarnitine (C4) − Energy
succinylcarnitine (C4)
1−(1−enyl−palmitoyl)−2−palmitoleoyl−GPC (P−16:0/16:1)*
X − 16944
(S)−3−hydroxybutyrylcarnitine
linolenoylcarnitine (C18:3)*
Phosphoethanolamine (PE) − Lipid
phosphocholine
glycerol 3−phosphate
prolylhydroxyproline
X − 14056
X − 18887
Taurolithocholate 3−sulfate − Lipid
tetradecanedioate (C14)
estriol 3−sulfate
3,5−dichloro−2,6−dihydroxybenzoic acid
X − 18838
hexadecenedioate (C16:1−DC)*

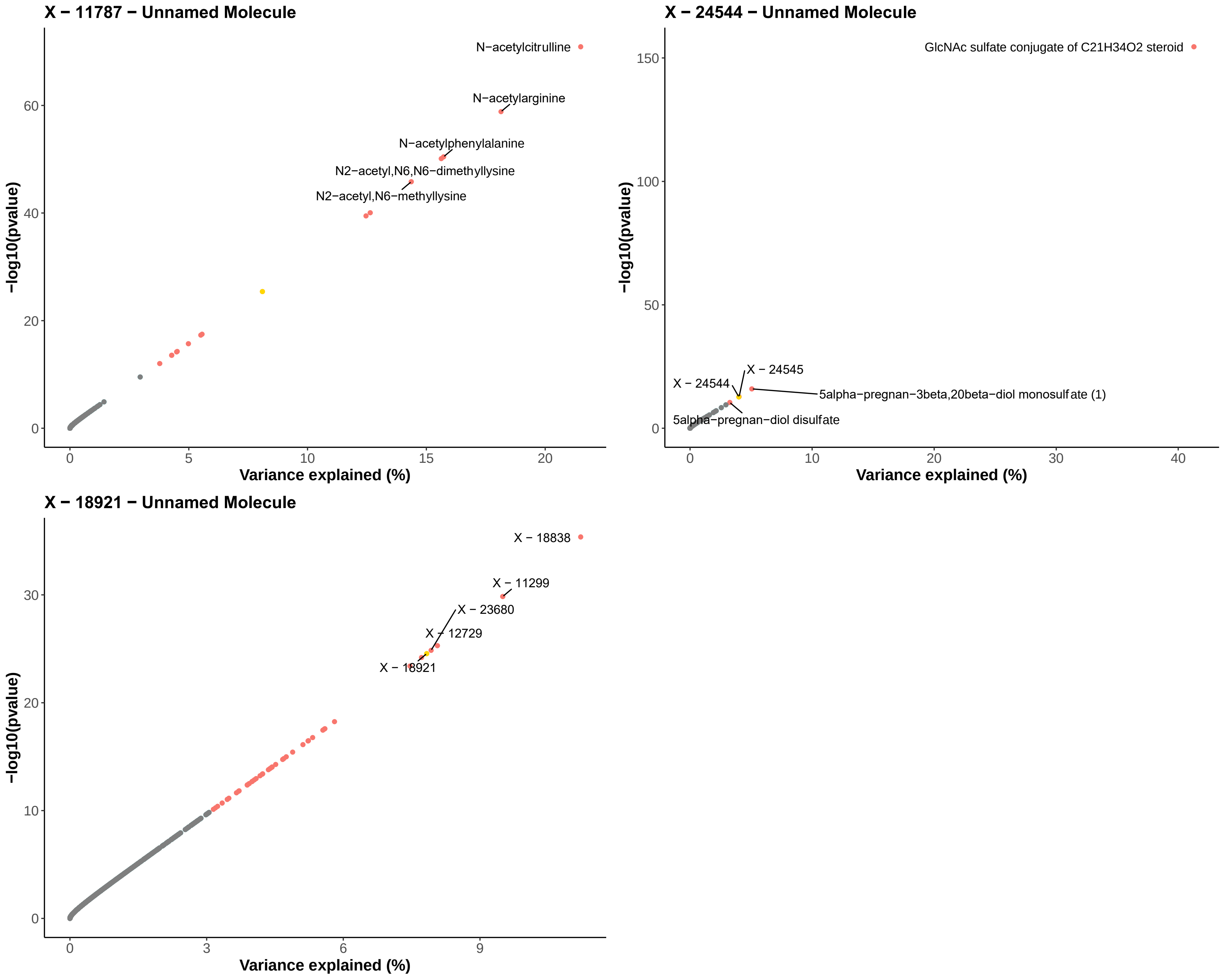

X − 11787 − Unnamed Molecule
N−acetylcitrulline
N−acetylarginine
N−acetylphenylalanine
N2−acetyl,N6,N6−dimethyllysine
N2−acetyl,N6−methyllysine
−log10(pvalue)
Variance explained (%)
X − 24544 − Unnamed Molecule
GlcNAc sulfate conjugate of C21H34O2 steroid
X − 24545
X − 24544
5alpha−pregnan−3beta,20beta−diol monosulfate (1)
5alpha−pregnan−diol disulfate
−log10(pvalue)
Variance explained (%)
X − 18921 − Unnamed Molecule
X − 18838
X − 11299
X − 23680
X − 12729
X − 18921
−log10(pvalue)
Variance explained (%)

**Figure S4.** Scatter plots of the variance explained by the weighted genetic risk score for each of the 27 metabolites included in MR analyses. Results are from linear regression analyses of GRS against all metabolites (exposure: GRS, outcome: metabolite) in BiB dataset 2 (see flowchart above for BiB dataset 2). The x-axes are $R^2$ expressed as a percentage and the y-axes are the -log10 P-value. The 5 metabolites with the lowest p-values in each scatter plot are labelled. Those with a -log10 P-value >10 are filled in red, with the remainder filled grey. The metabolite we are trying to proxy with the GRS is filled with gold. A highly specific GRS for a given metabolite would produce a scatter plot with the gold-filled point in the top right corner (high variance explained and strongly associated) with the remainder of the metabolites lower down towards the left corner.
